# Rapid spread of a SARS-CoV-2 Delta variant with a frameshift deletion in ORF7a

**DOI:** 10.1101/2021.08.18.21262089

**Authors:** Charles S.P. Foster, William D. Rawlinson

## Abstract

Australia is currently experiencing COVID-19 outbreaks from infection with SARS-CoV-2 Delta variants (B.1.617.2, AY.3). Analysis of the index case reveals a sub-consensus level of sequencing reads (∼25%) that support a 17-nucleotide deletion in ORF7a (ORF7a^Δ17del^). ORF7a^Δ17del^ induces a frameshift mutation in ORF7a, which truncates the peptide and potentially leads to reduced suppression of host restriction factor BST-2/CD317/Tetherin. Despite this, the mutation has rapidly become represented at the consensus level in subsequent cases: approximately 72% of SARS-CoV-2 genomes in the Australian outbreak possess ORF7a^Δ17del^, and 99.7% (1534/1538) of Delta genomes on GISAID with ORF7a^Δ17del^ originate from the current Australian outbreak (5 August 2021). The global abundance of this mutation might be underestimated given the difficulty of variant calling software correctly calling insertion/deletions (indels), the common inability of phylogenetics software to take indels into account, and the tendency of GISAID to not release submissions that contain a frameshift mutation (unless specifically requested). Overall, the rapid increase of persistent ORF7a^Δ17del^ variants is concerning, and suggests either a chance founder effect with a neutral mutation yet to be purged, or that the ORF7a^Δ17del^ mutation provides a direct selective advantage.

## Main text

SARS-CoV-2 encodes proteins that modulate antiviral responses (ORF3b, ORF6, ORF7a), including reducing type 1 interferon^1^. The ORF7a of SARS-CoV-2 is an ortholog of the corresponding SARS-CoV antagonist of host restriction factor BST-2/CD317/Tetherin that induces apoptosis^2,3^. Cells with reduced BST2 enhance SARS-CoV-2 replication thereby releasing more virus^3^. Deletion mutations at the SARS-CoV-2 C-terminus of ORF7a are frequent. An 81-nucleotide in-frame deletion in ORF7a removes signal peptide and beta strand sequences, which was detected in March 2020^2^. However, ORF7a deletions are generally transient. This may be due to reduced viral fitness, possibly as a consequence of reduced suppression of host innate immunity^1^.

Australia is currently experiencing COVID-19 outbreaks (>5000 cases) from infection with SARS-CoV-2 Delta variants (B.1.617.2, AY.3). These are characterised by increased frequency of detection of Delta variants with a 17-nt frameshift deletion in ORF7a (ORF7a^Δ17del^) and minimal other changes in the gene. This deletion spans genome positions 27607–27623 and leads to a truncated (78 vs 121 amino acids) peptide (Fig. 1A,B). The first virus of the current outbreak was collected on June 16, and the consensus genome exhibited a 100% match to a USA sequence with complete ORF7a (lacking ORF7a^Δ17del^) that was the likely progenitor (Fig. 1A). Notably, the index case of current Australian COVID-19 outbreaks and close contacts possessed a sub-consensus fraction of sequencing reads (∼25%) with ORF7a^Δ17del^. We sequenced the first Delta genome with consensus-level ORF7a^Δ17del^ in Australia on June 18, and ∼72% of SARS-CoV-2 genomes in the Australian outbreak possess ORF7a^Δ17del^ (Fig. 1C). This is likely an underestimate as it includes returned travellers and incomplete genome sequences. Analysis of 392,734 whole-genome SARS-CoV-2 Delta variant sequences from GISAID (5 August 2021) showed 99.7% (1534/1538) originate from the current Australian outbreak.

**Figure 1.**
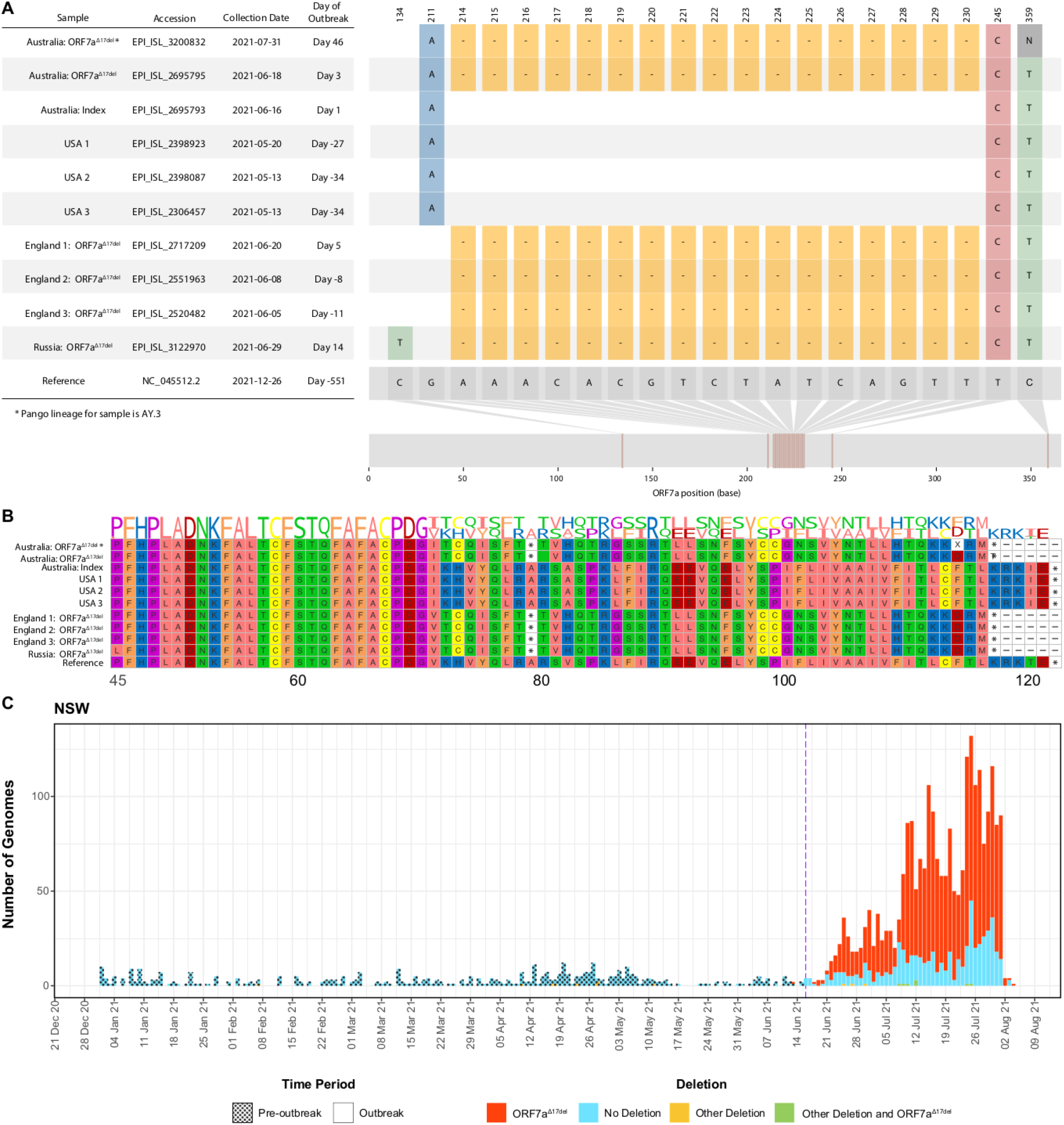
Visualisation of a 17-nucleotide deletion in ORF7a of Delta variant SARS-CoV-2 sequences (ORF7a ^Δ17del^) from Australia, 2021, the possible functional consequences at the peptide level, and the increasing frequency in NSW, Australia. Sequences chosen for visualisation include the index case of the current outbreaks (lacking ORF7a^Δ17del^), the first Australian case possessing ORF7a^Δ17del^, the only sample assigned to lineage AY.3 that possesses ORF7a ^17del^, the three USA samples (all lacking ORF7a^Δ17del^) that match the Australian index case, and the only four non-Australian samples to possess ORF7a^Δ17del^. Accession numbers for all samples analysed for this panel are in the Supplementary Appendix, available with the full text of this letter online. **Panel A** shows the sample dates of all chosen samples relative to the index case of the current outbreaks (16 June 2021), and all variants in the sample genomes relative to the SARS-CoV-2 reference genome (created in part using *snipit* (https://github.com/aineniamh/snipit). **Panel B** illustrates the consequences of ORF7a^Δ17del^ on the amino acid translation of ORF7a, leading to a premature stop codon after amino acid 78 and a truncated peptide sequence (∼64% complete). Amino acids 1–44 are identical to the SARS-CoV-2 reference genome in all samples and are not presented. **Panel C** demonstrates the number of genomes of all lineages from Australia from 01 January 2021 until 11 August 2021 with ORF7a^Δ17del^, ORF7a^Δ17del^ plus other deletion(s), other deletions in ORF7a only, or no deletions in ORF7a. The purple dashed line indicates the beginning of the current outbreaks in Australia, with shading also indicating pre-outbreak and outbreak samples.

The occurrence of ORF7a deletions in viruses from multiple countries throughout the pandemic suggests many deletions do not impair fitness, although gene truncation is generally associated with production of proteins with different or no activity. If ORF7a reduces BST2 effects in coronaviruses and BST2 knockdown enhances virion production^4^, the persistence of ORF7a^Δ17del^ suggests it arose by either a chance founder effect or provides a direct selective advantage compensating for loss of BST2 suppression. The latter should be investigated with functional studies of viruses containing ORF7a^Δ17del^, such as replicon and *in vitro* organoid research into phenotypic change, as done with S-protein mutations^5^. Given the many Delta variant COVID19 cases worldwide, the possible global spread of ORF7a^Δ17del^ should be closely monitored.

## Data Availability

All data were sourced from the NCBI and GISAID databases, and originate from submissions that were generated (a) in part by the authors, or (b) by authors from other institutions. All appropriate attributions are given following GISAID guidlines in the Supplemental Appendix, including Supplemental Table S1.

