## Supplementary Table S1 for "Rapid spread of a SARS-CoV-2 Delta variant with a frameshift deletion in ORF7a"

We gratefully acknowledge the following Authors from the Originating laboratories responsible for obtaining the specimens, as well as the Submitting laboratories where the genome data were generated and shared via GISAID, on which this research is based.

All Submitters of data may be contacted directly via www.gisaid.org

Authors are sorted alphabetically.

| Accession ID | Originating Laboratory | Submitting Laboratory | Authors |
|---|---|---|---|
| EPI_ISL_806726, EPI_ISL_2754125, EPI_ISL_2811990, EPI_ISL_2811993, EPI_ISL_2811994, EPI_ISL_2812818, EPI_ISL_2812852, EPI_ISL_2812858, EPI_ISL_2812873, EPI_ISL_2812900, EPI_ISL_2812911, EPI_ISL_2828076, EPI_ISL_2876027, EPI_ISL_2876032, EPI_ISL_2876033, EPI_ISL_2894135, EPI_ISL_3007298, EPI_ISL_3015128, EPI_ISL_3015148, EPI_ISL_3015150, EPI_ISL_3119466, EPI_ISL_3184881, EPI_ISL_3184899, EPI_ISL_3184905, EPI_ISL_3184937, EPI_ISL_3184941, EPI_ISL_3184944, EPI_ISL_3184997, EPI_ISL_3184998, EPI_ISL_3184999, EPI_ISL_3185000, EPI_ISL_3185001, EPI_ISL_3185002, EPI_ISL_3185053, EPI_ISL_3185103, EPI_ISL_3185104, EPI_ISL_3185105, EPI_ISL_3185106, EPI_ISL_3185107, EPI_ISL_3185108, EPI_ISL_3185109, EPI_ISL_3185110, EPI_ISL_3185111, EPI_ISL_3185112, EPI_ISL_3185163, EPI_ISL_3185164, EPI_ISL_3185165, EPI_ISL_3185166, EPI_ISL_3185167, EPI_ISL_3185168, EPI_ISL_3185169, EPI_ISL_3185170, EPI_ISL_3186076, EPI_ISL_3186080, EPI_ISL_3186082, EPI_ISL_3186088, EPI_ISL_3186089, EPI_ISL_3186090, EPI_ISL_3186181, EPI_ISL_3186182, EPI_ISL_3186238, EPI_ISL_3186239, EPI_ISL_3186244, EPI_ISL_3186245, EPI_ISL_3186246, EPI_ISL_3186247, EPI_ISL_3186254, EPI_ISL_3186255, EPI_ISL_3186256, EPI_ISL_3186271, EPI_ISL_3186273, EPI_ISL_3186465, EPI_ISL_3186624, EPI_ISL_3186625, EPI_ISL_3186626, EPI_ISL_3186627, EPI_ISL_3186628, EPI_ISL_3186629, EPI_ISL_3186630, EPI_ISL_3186631, EPI_ISL_3186632, EPI_ISL_3186633, EPI_ISL_3186659, EPI_ISL_3186694, EPI_ISL_3186695, EPI_ISL_3186724, EPI_ISL_3186736, EPI_ISL_3186836, EPI_ISL_3186837, EPI_ISL_3186838, EPI_ISL_3186839, EPI_ISL_3186840, EPI_ISL_3186841, EPI_ISL_3186842, EPI_ISL_3186843, EPI_ISL_3186866, EPI_ISL_3186867, EPI_ISL_3186868, EPI_ISL_3186869, EPI_ISL_3186870, EPI_ISL_3186871, EPI_ISL_3186872, EPI_ISL_3186873, EPI_ISL_3186874, EPI_ISL_3186952, EPI_ISL_3186953, EPI_ISL_3186954, EPI_ISL_3186955, EPI_ISL_3186956, EPI_ISL_3186957, EPI_ISL_3186958, EPI_ISL_3186959, EPI_ISL_3186960, EPI_ISL_3186961, EPI_ISL_3186962, EPI_ISL_3186963, EPI_ISL_3186964, EPI_ISL_3186965, EPI_ISL_3186966, EPI_ISL_3186967, EPI_ISL_3186968, EPI_ISL_3186969, EPI_ISL_3186970, EPI_ISL_3187045, EPI_ISL_3187046, EPI_ISL_3187047, EPI_ISL_3187048, EPI_ISL_3187049, EPI_ISL_3187050, EPI_ISL_3187051, EPI_ISL_3187052, EPI_ISL_3187053, EPI_ISL_3187054, EPI_ISL_3187055, EPI_ISL_3187056, EPI_ISL_3187057, EPI_ISL_3187058, EPI_ISL_3187059, EPI_ISL_3187060, EPI_ISL_3187061, EPI_ISL_3187062, EPI_ISL_3187063, EPI_ISL_3187064, EPI_ISL_3187115, EPI_ISL_3187116, EPI_ISL_3187117, EPI_ISL_3187118, EPI_ISL_3187119, EPI_ISL_3187120, EPI_ISL_3187121, EPI_ISL_3187122, EPI_ISL_3187123, EPI_ISL_3187124, EPI_ISL_3187125, EPI_ISL_3187126, EPI_ISL_3187127, EPI_ISL_3187128, EPI_ISL_3187129, EPI_ISL_3187130, EPI_ISL_3187131, EPI_ISL_3187215, EPI_ISL_3281055, EPI_ISL_3281162, EPI_ISL_3281163, EPI_ISL_3281164, EPI_ISL_3281165, EPI_ISL_3281166, EPI_ISL_3281167, EPI_ISL_3281168, EPI_ISL_3281169, EPI_ISL_3281170, EPI_ISL_3281171, EPI_ISL_3281172, EPI_ISL_3281173, EPI_ISL_3281209, EPI_ISL_3281210, EPI_ISL_3281211, EPI_ISL_3281212, EPI_ISL_3281213, EPI_ISL_3281214, EPI_ISL_3281215, EPI_ISL_3281216, EPI_ISL_3281217, EPI_ISL_3281218, EPI_ISL_3281219, EPI_ISL_3281220, EPI_ISL_3281221, EPI_ISL_3281222, EPI_ISL_3281223, EPI_ISL_3281224, EPI_ISL_3281225, EPI_ISL_3281285, EPI_ISL_3281286, EPI_ISL_3281287, EPI_ISL_3281288, EPI_ISL_3281289, EPI_ISL_3281290, EPI_ISL_3281291, EPI_ISL_3281292, EPI_ISL_3281293, EPI_ISL_3281294, EPI_ISL_3281295, EPI_ISL_3281296, EPI_ISL_3281297, EPI_ISL_3281298, EPI_ISL_3281299, EPI_ISL_3281300, EPI_ISL_3281301, EPI_ISL_3281302, EPI_ISL_3305419, EPI_ISL_3398402, EPI_ISL_3398409, EPI_ISL_3398415, EPI_ISL_3398421, EPI_ISL_3398422, EPI_ISL_3398423, EPI_ISL_3398437, EPI_ISL_3398454, EPI_ISL_3398455, EPI_ISL_3398483, EPI_ISL_3398508, EPI_ISL_3398509, EPI_ISL_3398518, EPI_ISL_3398533, EPI_ISL_3398543, EPI_ISL_3398544, EPI_ISL_3398545, EPI_ISL_3398546, EPI_ISL_3398552, EPI_ISL_3398553, EPI_ISL_3398566, EPI_ISL_3398567, EPI_ISL_3398568, EPI_ISL_3398581, EPI_ISL_3398582, EPI_ISL_3398583, EPI_ISL_3398584, EPI_ISL_3398585, EPI_ISL_3398593, EPI_ISL_3398594, EPI_ISL_3398595, EPI_ISL_3398609, EPI_ISL_3398610, EPI_ISL_3398611, EPI_ISL_3398646, EPI_ISL_3398647, EPI_ISL_3398660, EPI_ISL_3398661, EPI_ISL_3398662, EPI_ISL_3398663, EPI_ISL_3398664, EPI_ISL_3398665, EPI_ISL_3398666, EPI_ISL_3398667, EPI_ISL_3398668, EPI_ISL_3398669, EPI_ISL_3398670, EPI_ISL_3398671, EPI_ISL_3398672, EPI_ISL_3398673, EPI_ISL_3398674, EPI_ISL_3398675, EPI_ISL_3398676, EPI_ISL_3398677, EPI_ISL_3401768, EPI_ISL_3401769, EPI_ISL_3401770, EPI_ISL_3401771, EPI_ISL_3401772, EPI_ISL_3401773, EPI_ISL_3401774, EPI_ISL_3401775, EPI_ISL_3401776, EPI_ISL_3401777, EPI_ISL_3401778, EPI_ISL_3401779, EPI_ISL_3401780, EPI_ISL_3401781, EPI_ISL_3401782 | | | |
| see above | 4Cyte Pathology | NSW Health Pathology - Institute of Clinical Pathology and Medical Research; Westmead Hospital; University of Sydney | Arnott A.; CIDM-PH et al.; Draper J.; Gall M.; Martinez E.; Rockett R.; Sintchenko V.; on behalf of ICPMR |
| EPI_ISL_3281239 | ACT Pathology | NSW Health Pathology - Institute of Clinical Pathology and Medical Research; Westmead Hospital; University of Sydney | Arnott A.; Draper J.; Gall M.; Martinez E.; Rockett R.; Sintchenko V.; on behalf of ICPMR |
| EPI_ISL_1170946, EPI_ISL_1170947, EPI_ISL_1208400, EPI_ISL_1208401, EPI_ISL_1208402, EPI_ISL_1807318 | ACT Pathology | Schwessinger Lab | Ashley Jones; Benjamin Schwessinger; Craig Kennedy; Karina Kennedy; Kevin Murray; Megan McDonald; Ming-Dao Chia; Robert Lanfear; Robyn N Hall |
| EPI_ISL_1483700 | Area of Virology, Serology and Virology Division (SAVID), New South Wales Health Pathology Randwick | Area of Virology, Serology and Virology Division (SAVID), New South Wales Health Pathology Randwick | Au, J.; Bull, R.; Deveson, I.; Foster, C.; Jean, T.; Rawlinson, W.; Ruiz Silva, M.; Van Haal, S. |
| EPI_ISL_775242, EPI_ISL_775245, EPI_ISL_794671, EPI_ISL_1205815, EPI_ISL_1240991, EPI_ISL_1240992, EPI_ISL_1255309, EPI_ISL_1255310 | | | |
| see above | Area of Virology, Serology and Virology Division (SAViD), New South Wales Health Pathology Randwick | Area of Virology, Serology and Virology Division (SAViD), New South Wales Health Pathology Randwick | Au, J.; Bull, R.; Deveson, I.; Foster, C.; Rawlinson, W.; Ruiz Silva, M.; Van Haal, S.; Van Hal, S. |
| EPI_ISL_1010705, EPI_ISL_1010706 | Area of Virology, Serology and Virology Division (SAViD), New South Wales Health Pathology Randwick | Microbiology RPAH | Au, J.; Bull, R.; Deveson, I.; Foster, C.; Rawlinson, W.; Ruiz Silva, M.; Van Hal, S. |
| EPI_ISL_812347, EPI_ISL_812359, EPI_ISL_1005536, EPI_ISL_1005537, EPI_ISL_1005538, EPI_ISL_1005539, EPI_ISL_1005540, EPI_ISL_1069080, EPI_ISL_1098798, EPI_ISL_1098799, EPI_ISL_1121031, EPI_ISL_1121974, EPI_ISL_1121975, EPI_ISL_1121976, EPI_ISL_1184504, EPI_ISL_1184505, EPI_ISL_1184506, EPI_ISL_1293047, EPI_ISL_1293048, EPI_ISL_1293049, EPI_ISL_1293050, EPI_ISL_1406433, EPI_ISL_1406435, EPI_ISL_1424504, EPI_ISL_1424505, EPI_ISL_1424506, EPI_ISL_1424507, EPI_ISL_1424508, EPI_ISL_1494718, EPI_ISL_1494719, EPI_ISL_1494720, EPI_ISL_1494721, EPI_ISL_1494722, EPI_ISL_1494723, EPI_ISL_1516747, EPI_ISL_1516748, EPI_ISL_1543924, EPI_ISL_1543925, EPI_ISL_1543926, EPI_ISL_1543927, EPI_ISL_1563679, EPI_ISL_1563680, EPI_ISL_1563681, EPI_ISL_1563682, EPI_ISL_1615591, EPI_ISL_1615592, EPI_ISL_1615593, EPI_ISL_1615594, EPI_ISL_1615595, EPI_ISL_1615596, EPI_ISL_1615597, EPI_ISL_1615598, EPI_ISL_1633347, EPI_ISL_1633348, EPI_ISL_1633349, EPI_ISL_1633350, EPI_ISL_1633351, EPI_ISL_1633352, EPI_ISL_1633353, EPI_ISL_1672850, EPI_ISL_1672851, EPI_ISL_1672867, EPI_ISL_1911180, EPI_ISL_1911181, EPI_ISL_1911182, EPI_ISL_1911183, EPI_ISL_1911184, EPI_ISL_1911185, EPI_ISL_1911186, EPI_ISL_1911187, EPI_ISL_1911188, EPI_ISL_1911189, EPI_ISL_1911190, EPI_ISL_1911191, EPI_ISL_1913085, EPI_ISL_2404984, EPI_ISL_2404985, EPI_ISL_2404986, EPI_ISL_2404987, EPI_ISL_2404988, EPI_ISL_2404989, EPI_ISL_2404990, EPI_ISL_2404991, EPI_ISL_2404992, EPI_ISL_2455220, EPI_ISL_2534584, EPI_ISL_2534585, EPI_ISL_2534586, EPI_ISL_2534587, EPI_ISL_2633506, EPI_ISL_2633511, EPI_ISL_2633515, EPI_ISL_2633520, EPI_ISL_2633525, EPI_ISL_2650011, EPI_ISL_2650012, EPI_ISL_2695791, EPI_ISL_2695792, EPI_ISL_2695793, EPI_ISL_2695794, EPI_ISL_2695795, EPI_ISL_2695796, EPI_ISL_2716413, EPI_ISL_2716414, EPI_ISL_2716415, EPI_ISL_2716416, EPI_ISL_2716417, EPI_ISL_2716418, EPI_ISL_2716419, EPI_ISL_2716420, EPI_ISL_2716421, EPI_ISL_2716422, EPI_ISL_2716423, EPI_ISL_3200823, EPI_ISL_3200824, EPI_ISL_3200825, EPI_ISL_3200826, EPI_ISL_3200827, EPI_ISL_3200828, EPI_ISL_3200829, EPI_ISL_3200830, EPI_ISL_3200831, EPI_ISL_3200832, EPI_ISL_3221225, EPI_ISL_3221226, EPI_ISL_3221227, EPI_ISL_3221228, EPI_ISL_3221229, EPI_ISL_3221230, EPI_ISL_3221231, EPI_ISL_3221232, EPI_ISL_3221233, EPI_ISL_3221234, EPI_ISL_3221235, EPI_ISL_3332886 | | | |
| see above | Area of Virology, Serology and Virology Division (SAViD), New South Wales Health Pathology Randwick | Virology Research Laboratory; Area of Virology, Serology and Virology Division (SAViD), New South Wales Health Pathology Randwick | Au, J.; Bull, R.; Deveson, I.; Foster, C.; Rawlinson, W.; Ruiz Silva, M.; Van Hal, S.; Wong, M. |
| EPI_ISL_2811997, EPI_ISL_2876031, EPI_ISL_3184923, EPI_ISL_3187020, EPI_ISL_3281039, EPI_ISL_3398587 | Austech Medical Laboratories | NSW Health Pathology - Institute of Clinical Pathology and Medical Research; Westmead Hospital; University of Sydney | Arnott A.; CIDM-PH et al.; Draper J.; Gall M.; Martinez E.; Rockett R.; Sintchenko V.; on behalf of ICPMR |
| EPI_ISL_2680981, EPI_ISL_2812825, EPI_ISL_3007292, EPI_ISL_3007293, EPI_ISL_3184954, EPI_ISL_3185009, EPI_ISL_3186230, EPI_ISL_3186231, EPI_ISL_3186232, EPI_ISL_3186236, EPI_ISL_3186290, EPI_ISL_3305405 | | | |
| see above | Australian Clinical Labs | NSW Health Pathology - Institute of Clinical Pathology and Medical Research; Westmead Hospital; University of Sydney | Arnott A.; CIDM-PH et al.; Draper J.; Gall M.; Martinez E.; Rockett R.; Sintchenko V.; on behalf of ICPMR |
| EPI_ISL_2812901, EPI_ISL_2812912, EPI_ISL_2894149, EPI_ISL_2894150, EPI_ISL_3015151, EPI_ISL_3184898, EPI_ISL_3184903, EPI_ISL_3184927, EPI_ISL_3185055, EPI_ISL_3185113, EPI_ISL_3185114, EPI_ISL_3185153, EPI_ISL_3185154, EPI_ISL_3185171, EPI_ISL_3185172, EPI_ISL_3185173, EPI_ISL_3185174, EPI_ISL_3185175, EPI_ISL_3185176, EPI_ISL_3185177, EPI_ISL_3186585, EPI_ISL_3186587, EPI_ISL_3186588, EPI_ISL_3186589, EPI_ISL_3186590, EPI_ISL_3186660, EPI_ISL_3186696, EPI_ISL_3186725, EPI_ISL_3186726, EPI_ISL_3186875, EPI_ISL_3186876, EPI_ISL_3186971, EPI_ISL_3187065, EPI_ISL_3187066, EPI_ISL_3187067, EPI_ISL_3187068, EPI_ISL_3187069, EPI_ISL_3187070, EPI_ISL_3187071, EPI_ISL_3187072, EPI_ISL_3187073, EPI_ISL_3187074, EPI_ISL_3187075, EPI_ISL_3187167, EPI_ISL_3187168, EPI_ISL_3187169, EPI_ISL_3187176, EPI_ISL_3187216, EPI_ISL_3187227, EPI_ISL_3281051, EPI_ISL_3281226, EPI_ISL_3281227, EPI_ISL_3281228, EPI_ISL_3281229, EPI_ISL_3281230, EPI_ISL_3281231, EPI_ISL_3281303, EPI_ISL_3281304, EPI_ISL_3281305, EPI_ISL_3305482, EPI_ISL_3305500, EPI_ISL_3305502, EPI_ISL_3398403, EPI_ISL_3398424, EPI_ISL_3398425, EPI_ISL_3398426, EPI_ISL_3398586, EPI_ISL_3398596, EPI_ISL_3398612, EPI_ISL_3398656, EPI_ISL_3398678, EPI_ISL_3398679, EPI_ISL_3398680, EPI_ISL_3398681, EPI_ISL_3398682, EPI_ISL_3398683, EPI_ISL_3398684, EPI_ISL_3398702, EPI_ISL_3398704, EPI_ISL_3398705, EPI_ISL_3398706, EPI_ISL_3398708 | | | |
| see above | Australian Clinical Labs (formerly Healthscope Pathology) | NSW Health Pathology - Institute of Clinical Pathology and Medical Research; Westmead Hospital; University of Sydney | Arnott A.; CIDM-PH et al.; Draper J.; Gall M.; Martinez E.; Rockett R.; Sintchenko V.; on behalf of ICPMR |
| EPI_ISL_3184972, EPI_ISL_3185008, EPI_ISL_3185211, EPI_ISL_3185212 | Childrens Hospital Westmead | NSW Health Pathology - Institute of Clinical Pathology and Medical Research; Westmead Hospital; University of Sydney | CIDM-PH et al. |
| EPI_ISL_779398, EPI_ISL_2754117, EPI_ISL_2754118, EPI_ISL_2754119, EPI_ISL_2786935, EPI_ISL_2812870, EPI_ISL_3015129, EPI_ISL_3184888, EPI_ISL_3184900, EPI_ISL_3184907, EPI_ISL_3184993, EPI_ISL_3186091, EPI_ISL_3186092, EPI_ISL_3186093, EPI_ISL_3186228, EPI_ISL_3186229, EPI_ISL_3186592, EPI_ISL_3186593, EPI_ISL_3186672, EPI_ISL_3186673, EPI_ISL_3186674, EPI_ISL_3186697, EPI_ISL_3186753, EPI_ISL_3186877, EPI_ISL_3186878, EPI_ISL_3186879, EPI_ISL_3186880, EPI_ISL_3186881, EPI_ISL_3186972, EPI_ISL_3281033, EPI_ISL_3281174, EPI_ISL_3281175, EPI_ISL_3281176, EPI_ISL_3281177, EPI_ISL_3281178, EPI_ISL_3281179, EPI_ISL_3281180, EPI_ISL_3281181, EPI_ISL_3281182, EPI_ISL_3281183, EPI_ISL_3305424, EPI_ISL_3305426, EPI_ISL_3305466, EPI_ISL_3305494, EPI_ISL_3398375, EPI_ISL_3398438, EPI_ISL_3398456, EPI_ISL_3398457, EPI_ISL_3398458, EPI_ISL_3398502, EPI_ISL_3398569, EPI_ISL_3398570, EPI_ISL_3398617, EPI_ISL_3398654, EPI_ISL_3398655, EPI_ISL_3398685, EPI_ISL_3398686, EPI_ISL_3398687, EPI_ISL_3398688, EPI_ISL_3398689, EPI_ISL_3398690 | | | |
| see above | Douglass Hanly Moir Pathology | NSW Health Pathology - Institute of Clinical Pathology and Medical Research; Westmead Hospital; University of Sydney | Arnott A.; CIDM-PH et al.; Draper J.; Gall M.; Martinez E.; Rockett R.; Sintchenko V.; on behalf of ICPMR |
| EPI_ISL_872580, EPI_ISL_2754122, EPI_ISL_2754126, EPI_ISL_2754127, EPI_ISL_2811991, EPI_ISL_2811992, EPI_ISL_2812815, EPI_ISL_2812824, EPI_ISL_2812828, EPI_ISL_2812829, EPI_ISL_2812830, EPI_ISL_2812847, EPI_ISL_2812848, EPI_ISL_2812849, EPI_ISL_2812850, EPI_ISL_2812851, EPI_ISL_2812856, EPI_ISL_2812871, EPI_ISL_2812882, EPI_ISL_2812883, EPI_ISL_2812886, EPI_ISL_2812887, EPI_ISL_2812888, EPI_ISL_2812904, EPI_ISL_2812905, EPI_ISL_2812906, EPI_ISL_2812907, EPI_ISL_2812908, EPI_ISL_2812909, EPI_ISL_2812910, EPI_ISL_2828057, EPI_ISL_2828080, EPI_ISL_2828081, EPI_ISL_2828082, EPI_ISL_2828083, EPI_ISL_2828084, EPI_ISL_2828090, EPI_ISL_2828091, EPI_ISL_2876029, EPI_ISL_2876030, EPI_ISL_2894138, EPI_ISL_2894143, EPI_ISL_2894144, EPI_ISL_3007280, EPI_ISL_3007296, EPI_ISL_3007302, EPI_ISL_3015115, EPI_ISL_3015116, EPI_ISL_3015117, EPI_ISL_3015118, EPI_ISL_3015119, EPI_ISL_3015120, EPI_ISL_3015121, EPI_ISL_3015122, EPI_ISL_3015123, EPI_ISL_3015141, EPI_ISL_3015142, EPI_ISL_3015143, EPI_ISL_3015144, EPI_ISL_3015145, EPI_ISL_3015146, EPI_ISL_3015170, EPI_ISL_3015171, EPI_ISL_3015172, EPI_ISL_3015173, EPI_ISL_3015174, EPI_ISL_3015175, EPI_ISL_3015176, EPI_ISL_3184883, EPI_ISL_3184885, EPI_ISL_3184889, EPI_ISL_3184892, EPI_ISL_3184894, EPI_ISL_3184896, EPI_ISL_3184906, EPI_ISL_3184908, EPI_ISL_3184910, EPI_ISL_3184912, EPI_ISL_3184913, EPI_ISL_3184920, EPI_ISL_3184921, EPI_ISL_3184930, EPI_ISL_3184932, EPI_ISL_3184938, EPI_ISL_3184939, EPI_ISL_3184940, EPI_ISL_3184942, EPI_ISL_3184945, EPI_ISL_3184946, EPI_ISL_3184951, EPI_ISL_3184953, EPI_ISL_3185010, EPI_ISL_3185011, EPI_ISL_3185012, EPI_ISL_3185013, EPI_ISL_3185014, EPI_ISL_3185015, EPI_ISL_3185016, EPI_ISL_3185017, EPI_ISL_3185018, EPI_ISL_3185019, EPI_ISL_3185020, EPI_ISL_3185021, EPI_ISL_3185022, EPI_ISL_3185023, EPI_ISL_3185024, EPI_ISL_3185025, EPI_ISL_3185026, EPI_ISL_3185027, EPI_ISL_3185028, EPI_ISL_3185029, EPI_ISL_3185030, EPI_ISL_3185031, EPI_ISL_3185032, EPI_ISL_3185033, EPI_ISL_3185034, EPI_ISL_3185035, EPI_ISL_3185036, EPI_ISL_3185037, EPI_ISL_3185038, EPI_ISL_3185039, EPI_ISL_3185049, EPI_ISL_3185052, EPI_ISL_3185054, EPI_ISL_3185058, EPI_ISL_3185059, EPI_ISL_3185060, EPI_ISL_3185062, EPI_ISL_3185063, EPI_ISL_3185140, EPI_ISL_3185141, EPI_ISL_3185142, EPI_ISL_3185143, EPI_ISL_3185144, EPI_ISL_3185145, EPI_ISL_3185146, EPI_ISL_3185147, EPI_ISL_3185148, EPI_ISL_3185149, EPI_ISL_3185150, EPI_ISL_3185151, EPI_ISL_3185152, EPI_ISL_3185155, EPI_ISL_3185156, EPI_ISL_3185157, EPI_ISL_3185158, EPI_ISL_3185159, EPI_ISL_3185160, EPI_ISL_3185161, EPI_ISL_3185162, EPI_ISL_3185204, EPI_ISL_3185205, EPI_ISL_3185206, EPI_ISL_3185207, EPI_ISL_3185208, EPI_ISL_3185209, EPI_ISL_3185227, EPI_ISL_3185228, EPI_ISL_3185229, EPI_ISL_3185230, EPI_ISL_3185231, EPI_ISL_3185232, EPI_ISL_3185243, EPI_ISL_3185244, EPI_ISL_3185245, EPI_ISL_3185257, EPI_ISL_3186075, EPI_ISL_3186077, EPI_ISL_3186085, EPI_ISL_3186176, EPI_ISL_3186177, EPI_ISL_3186178, EPI_ISL_3186179, EPI_ISL_3186180, EPI_ISL_3186203, EPI_ISL_3186204, EPI_ISL_3186205, EPI_ISL_3186206, EPI_ISL_3186207, EPI_ISL_3186208, EPI_ISL_3186209, EPI_ISL_3186210, EPI_ISL_3186211, EPI_ISL_3186212, EPI_ISL_3186213, EPI_ISL_3186248, EPI_ISL_3186249, EPI_ISL_3186250, EPI_ISL_3186251, EPI_ISL_3186252, EPI_ISL_3186253, EPI_ISL_3186262, EPI_ISL_3186263, EPI_ISL_3186264, EPI_ISL_3186265, EPI_ISL_3186266, EPI_ISL_3186267, EPI_ISL_3186268, EPI_ISL_3186269, EPI_ISL_3186270, EPI_ISL_3186276, EPI_ISL_3186277, EPI_ISL_3186278, EPI_ISL_3186279, EPI_ISL_3186280, EPI_ISL_3186283, EPI_ISL_3186287, EPI_ISL_3186288, EPI_ISL_3186289, EPI_ISL_3186459, EPI_ISL_3186460, EPI_ISL_3186461, EPI_ISL_3186462, EPI_ISL_3186463, EPI_ISL_3186464, EPI_ISL_3186484, EPI_ISL_3186485, EPI_ISL_3186486, EPI_ISL_3186487, EPI_ISL_3186574, EPI_ISL_3186577, EPI_ISL_3186578, EPI_ISL_3186586, EPI_ISL_3186653, EPI_ISL_3186654, EPI_ISL_3186655, EPI_ISL_3186656, EPI_ISL_3186657, EPI_ISL_3186658, EPI_ISL_3186687, EPI_ISL_3186688, EPI_ISL_3186689, EPI_ISL_3186690, EPI_ISL_3186691, EPI_ISL_3186692, EPI_ISL_3186693, EPI_ISL_3186722, EPI_ISL_3186723, EPI_ISL_3186766, EPI_ISL_3186767, EPI_ISL_3186768, EPI_ISL_3186769, EPI_ISL_3186770, EPI_ISL_3186771, EPI_ISL_3186772, EPI_ISL_3186773, EPI_ISL_3186774, EPI_ISL_3186775, EPI_ISL_3186776, EPI_ISL_3186777, EPI_ISL_3186778, EPI_ISL_3186779, EPI_ISL_3186780, EPI_ISL_3186781, EPI_ISL_3186782, EPI_ISL_3186783, EPI_ISL_3186784, EPI_ISL_3186785, EPI_ISL_3186786, EPI_ISL_3186787, EPI_ISL_3186788, EPI_ISL_3186789, EPI_ISL_3186790, EPI_ISL_3186791, EPI_ISL_3186792, EPI_ISL_3186793, EPI_ISL_3186794, EPI_ISL_3186795, EPI_ISL_3186796, EPI_ISL_3186797, EPI_ISL_3186798, EPI_ISL_3186799, EPI_ISL_3186800, EPI_ISL_3186801, EPI_ISL_3186803, EPI_ISL_3186804, EPI_ISL_3186805, EPI_ISL_3186806, EPI_ISL_3186807, EPI_ISL_3186808, EPI_ISL_3186809, EPI_ISL_3186810, EPI_ISL_3186811, EPI_ISL_3186812, EPI_ISL_3186813, EPI_ISL_3186814, EPI_ISL_3186815, EPI_ISL_3186816, EPI_ISL_3186817, EPI_ISL_3186818, EPI_ISL_3186819, EPI_ISL_3186820, EPI_ISL_3186821, EPI_ISL_3186822, EPI_ISL_3186823, EPI_ISL_3186824, EPI_ISL_3186825, EPI_ISL_3186826, EPI_ISL_3186827, EPI_ISL_3186828, EPI_ISL_3186829, EPI_ISL_3186830, EPI_ISL_3186831, EPI_ISL_3186832, EPI_ISL_3186833, EPI_ISL_3186834, EPI_ISL_3186835, EPI_ISL_3186933, EPI_ISL_3186934, EPI_ISL_3186935, EPI_ISL_3186936, EPI_ISL_3186937, EPI_ISL_3186938, EPI_ISL_3186939, EPI_ISL_3186940, EPI_ISL_3186941, EPI_ISL_3186942, EPI_ISL_3186943, EPI_ISL_3186944, EPI_ISL_3186945, EPI_ISL_3186946, EPI_ISL_3186947, EPI_ISL_3186948, EPI_ISL_3186949, EPI_ISL_3186950, EPI_ISL_3187021, EPI_ISL_3187022, EPI_ISL_3187023, EPI_ISL_3187024, EPI_ISL_3187025, EPI_ISL_3187026, EPI_ISL_3187027, EPI_ISL_3187028, EPI_ISL_3187029, EPI_ISL_3187030, EPI_ISL_3187031, EPI_ISL_3187032, EPI_ISL_3187033, EPI_ISL_3187034, EPI_ISL_3187035, EPI_ISL_3187036, EPI_ISL_3187037, EPI_ISL_3187038, EPI_ISL_3187039, EPI_ISL_3187040, EPI_ISL_3187041, EPI_ISL_3187042, EPI_ISL_3187043, EPI_ISL_3187044, EPI_ISL_3187109, EPI_ISL_3187110, EPI_ISL_3187111, EPI_ISL_3187112, EPI_ISL_3187113, EPI_ISL_3187114, EPI_ISL_3187213, EPI_ISL_3187214, EPI_ISL_3187219, EPI_ISL_3187220, EPI_ISL_3187229, EPI_ISL_3281019, EPI_ISL_3281022, EPI_ISL_3281024, EPI_ISL_3281025, EPI_ISL_3281031, EPI_ISL_3281044, EPI_ISL_3281045, EPI_ISL_3281046, EPI_ISL_3281054, EPI_ISL_3281060, EPI_ISL_3281061, EPI_ISL_3281062, EPI_ISL_3281063, EPI_ISL_3281064, EPI_ISL_3281065, EPI_ISL_3281066, EPI_ISL_3281067, EPI_ISL_3281068, EPI_ISL_3281069, EPI_ISL_3281070, EPI_ISL_3281071, EPI_ISL_3281072, EPI_ISL_3281073, EPI_ISL_3281074, EPI_ISL_3281075, EPI_ISL_3281076, EPI_ISL_3281077, EPI_ISL_3281078, EPI_ISL_3281079, EPI_ISL_3281080, EPI_ISL_3281081, EPI_ISL_3281082, EPI_ISL_3281083, EPI_ISL_3281084, EPI_ISL_3281085, EPI_ISL_3281086, EPI_ISL_3281087, EPI_ISL_3281088, EPI_ISL_3281089, EPI_ISL_3281090, EPI_ISL_3281091, EPI_ISL_3281092, EPI_ISL_3281093, EPI_ISL_3281094, EPI_ISL_3281095, EPI_ISL_3281096, EPI_ISL_3281097, EPI_ISL_3281098, EPI_ISL_3281099, EPI_ISL_3281100, EPI_ISL_3281101, EPI_ISL_3281102, EPI_ISL_3281103, EPI_ISL_3281104, EPI_ISL_3281105, EPI_ISL_3281106, EPI_ISL_3281107, EPI_ISL_3281108, EPI_ISL_3281109, EPI_ISL_3281110, EPI_ISL_3281111, EPI_ISL_3281112, EPI_ISL_3281113, EPI_ISL_3281114, EPI_ISL_3281115, EPI_ISL_3281116, EPI_ISL_3281117, EPI_ISL_3281118, EPI_ISL_3281119, EPI_ISL_3281120, EPI_ISL_3281121, EPI_ISL_3281122, EPI_ISL_3281123, EPI_ISL_3281124, EPI_ISL_3281125, EPI_ISL_3281126, EPI_ISL_3281127, EPI_ISL_3281128, EPI_ISL_3281129, EPI_ISL_3281130, EPI_ISL_3281131, EPI_ISL_3281132, EPI_ISL_3281133, EPI_ISL_3281134, EPI_ISL_3281135, EPI_ISL_3281136, EPI_ISL_3281242, EPI_ISL_3281243, EPI_ISL_3281244, EPI_ISL_3281245, EPI_ISL_3281246, EPI_ISL_3281247, EPI_ISL_3281248, EPI_ISL_3281249, EPI_ISL_3281250, EPI_ISL_3281251, EPI_ISL_3281252, EPI_ISL_3281253, EPI_ISL_3281254, EPI_ISL_3281255, EPI_ISL_3281256, EPI_ISL_3281257, EPI_ISL_3281258, EPI_ISL_3281259, EPI_ISL_3281260, EPI_ISL_3281261, EPI_ISL_3281262, EPI_ISL_3281263, EPI_ISL_3281264, EPI_ISL_3281265, EPI_ISL_3281266, EPI_ISL_3281267, EPI_ISL_3281268, EPI_ISL_3281269, EPI_ISL_3281270, EPI_ISL_3281271, EPI_ISL_3281272, EPI_ISL_3281273, EPI_ISL_3281274, EPI_ISL_3281275, EPI_ISL_3281276, EPI_ISL_3281277, EPI_ISL_3281278, EPI_ISL_3281279, EPI_ISL_3281280, EPI_ISL_3281281, EPI_ISL_3281282, EPI_ISL_3281283, EPI_ISL_3281284, EPI_ISL_3281341, EPI_ISL_3281342, EPI_ISL_3281343, EPI_ISL_3281344, EPI_ISL_3281345, EPI_ISL_3281346, EPI_ISL_3281347, EPI_ISL_3281348, EPI_ISL_3281349, EPI_ISL_3281350, EPI_ISL_3281351, EPI_ISL_3281352, EPI_ISL_3281353, EPI_ISL_3281354, EPI_ISL_3281355, EPI_ISL_3281356, EPI_ISL_3281357, EPI_ISL_3281358, EPI_ISL_3281359, EPI_ISL_3281360, EPI_ISL_3281361, EPI_ISL_3281362, EPI_ISL_3281363, EPI_ISL_3281364, EPI_ISL_3281365, EPI_ISL_3281366, EPI_ISL_3281368, EPI_ISL_3281369, EPI_ISL_3281370, EPI_ISL_3281371, EPI_ISL_3281372, EPI_ISL_3281373, EPI_ISL_3281374, EPI_ISL_3281375, EPI_ISL_3281376, EPI_ISL_3281377, EPI_ISL_3281378, EPI_ISL_3281379, EPI_ISL_3281380, EPI_ISL_3281381, EPI_ISL_3281382, EPI_ISL_3281383, EPI_ISL_3281384, EPI_ISL_3281385, EPI_ISL_3281386, EPI_ISL_3281387, EPI_ISL_3281388, EPI_ISL_3281389, EPI_ISL_3281390, EPI_ISL_3281391, EPI_ISL_3281392, EPI_ISL_3281393, EPI_ISL_3281394, EPI_ISL_3281395, EPI_ISL_3305417, EPI_ISL_3305441, EPI_ISL_3305451, EPI_ISL_3305453, EPI_ISL_3305454, EPI_ISL_3305464, EPI_ISL_3305468, EPI_ISL_3305469, EPI_ISL_3305470, EPI_ISL_3305515, EPI_ISL_3305517, EPI_ISL_3305518, EPI_ISL_3305520, EPI_ISL_3305522, EPI_ISL_3305524, EPI_ISL_3305526, EPI_ISL_3305528, EPI_ISL_3305529, EPI_ISL_3305531, EPI_ISL_3305533, EPI_ISL_3305535, EPI_ISL_3305537, EPI_ISL_3305539, EPI_ISL_3305541, EPI_ISL_3305542, EPI_ISL_3398377, EPI_ISL_3398378, EPI_ISL_3398379, EPI_ISL_3398380, EPI_ISL_3398386, EPI_ISL_3398392, EPI_ISL_3398393, EPI_ISL_3398408, EPI_ISL_3398416, EPI_ISL_3398417, EPI_ISL_3398418, EPI_ISL_3398419, EPI_ISL_3398420, EPI_ISL_3398430, EPI_ISL_3398431, EPI_ISL_3398432, EPI_ISL_3398433, EPI_ISL_3398434, EPI_ISL_3398435, EPI_ISL_3398436, EPI_ISL_3398451, EPI_ISL_3398452, EPI_ISL_3398453, EPI_ISL_3398473, EPI_ISL_3398474, EPI_ISL_3398475, EPI_ISL_3398476, EPI_ISL_3398477, EPI_ISL_3398478, EPI_ISL_3398479, EPI_ISL_3398489, EPI_ISL_3398490, EPI_ISL_3398491, EPI_ISL_3398504, EPI_ISL_3398505, EPI_ISL_3398506, EPI_ISL_3398507, EPI_ISL_3398510, EPI_ISL_3398511, EPI_ISL_3398512, EPI_ISL_3398513, EPI_ISL_3398514, EPI_ISL_3398521, EPI_ISL_3398522, EPI_ISL_3398523, EPI_ISL_3398524, EPI_ISL_3398525, EPI_ISL_3398526, EPI_ISL_3398527, EPI_ISL_3398528, EPI_ISL_3398529, EPI_ISL_3398530, EPI_ISL_3398531, EPI_ISL_3398532, EPI_ISL_3398549, EPI_ISL_3398550, EPI_ISL_3398551, EPI_ISL_3398559, EPI_ISL_3398560, EPI_ISL_3398574, EPI_ISL_3398575, EPI_ISL_3398576, EPI_ISL_3398577, EPI_ISL_3398578, EPI_ISL_3398579, EPI_ISL_3398580, EPI_ISL_3398590, EPI_ISL_3398591, EPI_ISL_3398592, EPI_ISL_3398602, EPI_ISL_3398603, EPI_ISL_3398604, EPI_ISL_3398605, EPI_ISL_3398606, EPI_ISL_3398607, EPI_ISL_3398608, EPI_ISL_3398622, EPI_ISL_3398623, EPI_ISL_3398624, EPI_ISL_3398625, EPI_ISL_3398626, EPI_ISL_3398627, EPI_ISL_3398628, EPI_ISL_3398629, EPI_ISL_3398630, EPI_ISL_3398631, EPI_ISL_3398632, EPI_ISL_3398633, EPI_ISL_3398634, EPI_ISL_3398635, EPI_ISL_3398636, EPI_ISL_3398637, EPI_ISL_3398638, EPI_ISL_3398639, EPI_ISL_3398640, EPI_ISL_3398641, EPI_ISL_3398642, EPI_ISL_3398643, EPI_ISL_3398644, EPI_ISL_3398645, EPI_ISL_3398727, EPI_ISL_3398728, EPI_ISL_3398729, EPI_ISL_3398730, EPI_ISL_3398731, EPI_ISL_3398732, EPI_ISL_3398733, EPI_ISL_3398734, EPI_ISL_3398735, EPI_ISL_3398736, EPI_ISL_3398737, EPI_ISL_3398738, EPI_ISL_3398739, EPI_ISL_3398740, EPI_ISL_3398741, EPI_ISL_3398742, EPI_ISL_3398743, EPI_ISL_3398744, EPI_ISL_3398745, EPI_ISL_3398746, EPI_ISL_3398747, EPI_ISL_3398748, EPI_ISL_3398749, EPI_ISL_3398750, EPI_ISL_3398751, EPI_ISL_3398752, EPI_ISL_3398753, EPI_ISL_3398754, EPI_ISL_3398755, EPI_ISL_3398756, EPI_ISL_3401670, EPI_ISL_3401671, EPI_ISL_3401672, EPI_ISL_3401673, EPI_ISL_3401674, EPI_ISL_3401675, EPI_ISL_3401676, EPI_ISL_3401677, EPI_ISL_3401678, EPI_ISL_3401679, EPI_ISL_3401680, EPI_ISL_3401681, EPI_ISL_3401682, EPI_ISL_3401683, EPI_ISL_3401684, EPI_ISL_3401793, EPI_ISL_3401794, EPI_ISL_3401795, EPI_ISL_3401796, EPI_ISL_3401797, EPI_ISL_3401798, EPI_ISL_3401799, EPI_ISL_3401800, EPI_ISL_3401801, EPI_ISL_3401802, EPI_ISL_3401803, EPI_ISL_3401804, EPI_ISL_3401805, EPI_ISL_3401806, EPI_ISL_3401807, EPI_ISL_3401808, EPI_ISL_3401809, EPI_ISL_3401810, EPI_ISL_3401811, EPI_ISL_3401812, EPI_ISL_3401813, EPI_ISL_3401814, EPI_ISL_3401815, EPI_ISL_3401816, EPI_ISL_3401817, EPI_ISL_3401818, EPI_ISL_3401819, EPI_ISL_3401820, EPI_ISL_3401821, EPI_ISL_3401822, EPI_ISL_3401823, EPI_ISL_3401824, EPI_ISL_3401825, EPI_ISL_3401826, EPI_ISL_3401827, EPI_ISL_3401828, EPI_ISL_3401829, EPI_ISL_3401830, EPI_ISL_3401831, EPI_ISL_3401832, EPI_ISL_3401833, EPI_ISL_3401834, EPI_ISL_3401835, EPI_ISL_3401836, EPI_ISL_3401837, EPI_ISL_3401838, EPI_ISL_3401839, EPI_ISL_3401840, EPI_ISL_3401841 | | | |
| see above | Histopath | NSW Health Pathology - Institute of Clinical Pathology and Medical Research; Westmead Hospital; University of Sydney | Arnott A.; CIDM-PH et al.; Draper J.; Gall M.; Martinez E.; Rockett R.; Sintchenko V.; on behalf of ICPMR |

| | | | |
|---|---|---|---|
| EPI_ISL_2828075, EPI_ISL_2828077, EPI_ISL_2828078, EPI_ISL_2828086, EPI_ISL_3398387, EPI_ISL_3398389, EPI_ISL_3398390, EPI_ISL_3398391 | | | |
| see above | Laverty | NSW Health Pathology - Institute of Clinical Pathology and Medical Research; Westmead Hospital; University of Sydney | Arnott A.; CIDM-PH et al.; Draper J.; Gall M.; Martinez E.; Rockett R.; Sintchenko V.; on behalf of ICPMR |
| EPI_ISL_767938, EPI_ISL_872573, EPI_ISL_872576, EPI_ISL_2650467, EPI_ISL_2680984, EPI_ISL_2680985, EPI_ISL_2754116, EPI_ISL_2754120, EPI_ISL_2754123, EPI_ISL_2754124, EPI_ISL_2786932, EPI_ISL_2811989, EPI_ISL_2811995, EPI_ISL_2812816, EPI_ISL_2812820, EPI_ISL_2812853, EPI_ISL_2812854, EPI_ISL_2812874, EPI_ISL_2812875, EPI_ISL_2812876, EPI_ISL_2812880, EPI_ISL_2812881, EPI_ISL_2812889, EPI_ISL_2812890, EPI_ISL_2812898, EPI_ISL_2812899, EPI_ISL_2812913, EPI_ISL_2812914, EPI_ISL_2812915, EPI_ISL_2812916, EPI_ISL_2812917, EPI_ISL_2861253, EPI_ISL_2861254, EPI_ISL_2876034, EPI_ISL_2876035, EPI_ISL_2876036, EPI_ISL_2876037, EPI_ISL_2877039, EPI_ISL_2877040, EPI_ISL_2877041, EPI_ISL_2877042, EPI_ISL_2894134, EPI_ISL_2894139, EPI_ISL_2894140, EPI_ISL_2894145, EPI_ISL_2894146, EPI_ISL_3007289, EPI_ISL_3007290, EPI_ISL_3007291, EPI_ISL_3007299, EPI_ISL_3007300, EPI_ISL_3015125, EPI_ISL_3015126, EPI_ISL_3015149, EPI_ISL_3015152, EPI_ISL_3015153, EPI_ISL_3015154, EPI_ISL_3015155, EPI_ISL_3015156, EPI_ISL_3015157, EPI_ISL_3119461, EPI_ISL_3119462, EPI_ISL_3119463, EPI_ISL_3184884, EPI_ISL_3184893, EPI_ISL_3184897, EPI_ISL_3184914, EPI_ISL_3184919, EPI_ISL_3184924, EPI_ISL_3184936, EPI_ISL_3184943, EPI_ISL_3184955, EPI_ISL_3184956, EPI_ISL_3184985, EPI_ISL_3184986, EPI_ISL_3184987, EPI_ISL_3184988, EPI_ISL_3184989, EPI_ISL_3184990, EPI_ISL_3184991, EPI_ISL_3184992, EPI_ISL_3184994, EPI_ISL_3184995, EPI_ISL_3184996, EPI_ISL_3185045, EPI_ISL_3185046, EPI_ISL_3185051, EPI_ISL_3185115, EPI_ISL_3185116, EPI_ISL_3185178, EPI_ISL_3185179, EPI_ISL_3185180, EPI_ISL_3185181, EPI_ISL_3185182, EPI_ISL_3185183, EPI_ISL_3185184, EPI_ISL_3185185, EPI_ISL_3185223, EPI_ISL_3185246, EPI_ISL_3186084, EPI_ISL_3186086, EPI_ISL_3186087, EPI_ISL_3186183, EPI_ISL_3186184, EPI_ISL_3186218, EPI_ISL_3186219, EPI_ISL_3186220, EPI_ISL_3186221, EPI_ISL_3186222, EPI_ISL_3186223, EPI_ISL_3186224, EPI_ISL_3186225, EPI_ISL_3186226, EPI_ISL_3186233, EPI_ISL_3186234, EPI_ISL_3186235, EPI_ISL_3186237, EPI_ISL_3186240, EPI_ISL_3186241, EPI_ISL_3186242, EPI_ISL_3186243, EPI_ISL_3186281, EPI_ISL_3186282, EPI_ISL_3186284, EPI_ISL_3186285, EPI_ISL_3186286, EPI_ISL_3186468, EPI_ISL_3186469, EPI_ISL_3186470, EPI_ISL_3186471, EPI_ISL_3186575, EPI_ISL_3186576, EPI_ISL_3186594, EPI_ISL_3186595, EPI_ISL_3186596, EPI_ISL_3186597, EPI_ISL_3186598, EPI_ISL_3186599, EPI_ISL_3186600, EPI_ISL_3186601, EPI_ISL_3186602, EPI_ISL_3186603, EPI_ISL_3186604, EPI_ISL_3186605, EPI_ISL_3186647, EPI_ISL_3186648, EPI_ISL_3186649, EPI_ISL_3186650, EPI_ISL_3186651, EPI_ISL_3186652, EPI_ISL_3186675, EPI_ISL_3186727, EPI_ISL_3186728, EPI_ISL_3186745, EPI_ISL_3186746, EPI_ISL_3186747, EPI_ISL_3186749, EPI_ISL_3186750, EPI_ISL_3186754, EPI_ISL_3186755, EPI_ISL_3186757, EPI_ISL_3186758, EPI_ISL_3186759, EPI_ISL_3186760, EPI_ISL_3186761, EPI_ISL_3186762, EPI_ISL_3186763, EPI_ISL_3186764, EPI_ISL_3186882, EPI_ISL_3186883, EPI_ISL_3186884, EPI_ISL_3186885, EPI_ISL_3186886, EPI_ISL_3186887, EPI_ISL_3186888, EPI_ISL_3186889, EPI_ISL_3186890, EPI_ISL_3186891, EPI_ISL_3186892, EPI_ISL_3186893, EPI_ISL_3186894, EPI_ISL_3186895, EPI_ISL_3186896, EPI_ISL_3186897, EPI_ISL_3187076, EPI_ISL_3187077, EPI_ISL_3187078, EPI_ISL_3187079, EPI_ISL_3187080, EPI_ISL_3187081, EPI_ISL_3187082, EPI_ISL_3187083, EPI_ISL_3187084, EPI_ISL_3187085, EPI_ISL_3187086, EPI_ISL_3187087, EPI_ISL_3187088, EPI_ISL_3187089, EPI_ISL_3187170, EPI_ISL_3187171, EPI_ISL_3187172, EPI_ISL_3187173, EPI_ISL_3187174, EPI_ISL_3187177, EPI_ISL_3187178, EPI_ISL_3187179, EPI_ISL_3187180, EPI_ISL_3187181, EPI_ISL_3187182, EPI_ISL_3187183, EPI_ISL_3187184, EPI_ISL_3187185, EPI_ISL_3187186, EPI_ISL_3187187, EPI_ISL_3187188, EPI_ISL_3187189, EPI_ISL_3187190, EPI_ISL_3187191, EPI_ISL_3187197, EPI_ISL_3187200, EPI_ISL_3187203, EPI_ISL_3187204, EPI_ISL_3187205, EPI_ISL_3187206, EPI_ISL_3187207, EPI_ISL_3187208, EPI_ISL_3187209, EPI_ISL_3187210, EPI_ISL_3187218, EPI_ISL_3187222, EPI_ISL_3187226, EPI_ISL_3281020, EPI_ISL_3281021, EPI_ISL_3281023, EPI_ISL_3281026, EPI_ISL_3281028, EPI_ISL_3281034, EPI_ISL_3281035, EPI_ISL_3281040, EPI_ISL_3281041, EPI_ISL_3281042, EPI_ISL_3281057, EPI_ISL_3281058, EPI_ISL_3281184, EPI_ISL_3281185, EPI_ISL_3281186, EPI_ISL_3281187, EPI_ISL_3281188, EPI_ISL_3281189, EPI_ISL_3281190, EPI_ISL_3281191, EPI_ISL_3281192, EPI_ISL_3281193, EPI_ISL_3281194, EPI_ISL_3281195, EPI_ISL_3281196, EPI_ISL_3281197, EPI_ISL_3281198, EPI_ISL_3281199, EPI_ISL_3281200, EPI_ISL_3281201, EPI_ISL_3281202, EPI_ISL_3281203, EPI_ISL_3281204, EPI_ISL_3281205, EPI_ISL_3281206, EPI_ISL_3281207, EPI_ISL_3281208, EPI_ISL_3281402, EPI_ISL_3281403, EPI_ISL_3281404, EPI_ISL_3281405, EPI_ISL_3281406, EPI_ISL_3281407, EPI_ISL_3281408, EPI_ISL_3305434, EPI_ISL_3305435, EPI_ISL_3305447, EPI_ISL_3305449, EPI_ISL_3305491, EPI_ISL_3305496, EPI_ISL_3305498, EPI_ISL_3398374, EPI_ISL_3398376, EPI_ISL_3398384, EPI_ISL_3398385, EPI_ISL_3398412, EPI_ISL_3398413, EPI_ISL_3398414, EPI_ISL_3398484, EPI_ISL_3398485, EPI_ISL_3398493, EPI_ISL_3398494, EPI_ISL_3398495, EPI_ISL_3398515, EPI_ISL_3398516, EPI_ISL_3398517, EPI_ISL_3398536, EPI_ISL_3398537, EPI_ISL_3398547, EPI_ISL_3398561, EPI_ISL_3398562, EPI_ISL_3398571, EPI_ISL_3398572, EPI_ISL_3398573, EPI_ISL_3398588, EPI_ISL_3398613, EPI_ISL_3398614, EPI_ISL_3398615, EPI_ISL_3398616, EPI_ISL_3398652, EPI_ISL_3398653, EPI_ISL_3398657, EPI_ISL_3398658, EPI_ISL_3398691, EPI_ISL_3398692, EPI_ISL_3398693, EPI_ISL_3398694, EPI_ISL_3398695, EPI_ISL_3398696, EPI_ISL_3398697, EPI_ISL_3398698, EPI_ISL_3398699, EPI_ISL_3398700, EPI_ISL_3398701, EPI_ISL_3401686, EPI_ISL_3401687, EPI_ISL_3401688, EPI_ISL_3401689, EPI_ISL_3401690, EPI_ISL_3401691, EPI_ISL_3401692, EPI_ISL_3401693, EPI_ISL_3401694, EPI_ISL_3401695, EPI_ISL_3401696, EPI_ISL_3401697, EPI_ISL_3401698, EPI_ISL_3401699, EPI_ISL_3401700, EPI_ISL_3401701, EPI_ISL_3401702, EPI_ISL_3401703, EPI_ISL_3401704, EPI_ISL_3401705, EPI_ISL_3401706, EPI_ISL_3401707, EPI_ISL_3401708, EPI_ISL_3401709, EPI_ISL_3401710, EPI_ISL_3401711, EPI_ISL_3401712, EPI_ISL_3401713, EPI_ISL_3401714, EPI_ISL_3401715, EPI_ISL_3401716 | | | |
| see above | Laverty Pathology | NSW Health Pathology - Institute of Clinical Pathology and Medical Research; Westmead Hospital; University of Sydney | Arnott A.; CIDM-PH et al.; Draper J.; Gall M.; Martinez E.; Rockett R.; Sintchenko V.; on behalf of ICPMR |
| EPI_ISL_2861250, EPI_ISL_2876028 | Liverpool Hospital | NSW Health Pathology - Institute of Clinical Pathology and Medical Research; Westmead Hospital; University of Sydney | CIDM-PH et al. |
| EPI_ISL_806725, EPI_ISL_845802, EPI_ISL_845803, EPI_ISL_845804, EPI_ISL_857309, EPI_ISL_2754128, EPI_ISL_2812884, EPI_ISL_2812903, EPI_ISL_2876026, EPI_ISL_2877043, EPI_ISL_2877044, EPI_ISL_2894136, EPI_ISL_2894147, EPI_ISL_3184890, EPI_ISL_3184909, EPI_ISL_3185044, EPI_ISL_3185050, EPI_ISL_3185061, EPI_ISL_3185186, EPI_ISL_3185187, EPI_ISL_3185188, EPI_ISL_3185189, EPI_ISL_3185190, EPI_ISL_3185191, EPI_ISL_3185192, EPI_ISL_3185247, EPI_ISL_3185248, EPI_ISL_3185249, EPI_ISL_3186214, EPI_ISL_3186215, EPI_ISL_3186216, EPI_ISL_3186257, EPI_ISL_3186258, EPI_ISL_3186613, EPI_ISL_3186634, EPI_ISL_3186635, EPI_ISL_3186636, EPI_ISL_3186637, EPI_ISL_3186638, EPI_ISL_3186661, EPI_ISL_3186662, EPI_ISL_3186663, EPI_ISL_3186664, EPI_ISL_3186665, EPI_ISL_3186666, EPI_ISL_3186667, EPI_ISL_3186668, EPI_ISL_3186669, EPI_ISL_3186670, EPI_ISL_3186705, EPI_ISL_3186706, EPI_ISL_3186707, EPI_ISL_3186708, EPI_ISL_3186709, EPI_ISL_3186710, EPI_ISL_3186711, EPI_ISL_3186712, EPI_ISL_3186713, EPI_ISL_3186714, EPI_ISL_3186720, EPI_ISL_3186721, EPI_ISL_3186729, EPI_ISL_3186730, EPI_ISL_3186731, EPI_ISL_3186732, EPI_ISL_3186733, EPI_ISL_3186734, EPI_ISL_3186735, EPI_ISL_3186737, EPI_ISL_3186738, EPI_ISL_3186739, EPI_ISL_3186740, EPI_ISL_3186741, EPI_ISL_3186742, EPI_ISL_3186844, EPI_ISL_3186845, EPI_ISL_3186846, EPI_ISL_3186847, EPI_ISL_3186848, EPI_ISL_3186849, EPI_ISL_3186850, EPI_ISL_3186851, EPI_ISL_3186852, EPI_ISL_3186853, EPI_ISL_3186854, EPI_ISL_3186855, EPI_ISL_3186856, EPI_ISL_3186857, EPI_ISL_3186858, EPI_ISL_3186859, EPI_ISL_3186860, EPI_ISL_3186861, EPI_ISL_3186862, EPI_ISL_3186863, EPI_ISL_3186864, EPI_ISL_3186865, EPI_ISL_3186996, EPI_ISL_3186997, EPI_ISL_3186998, EPI_ISL_3187094, EPI_ISL_3187095, EPI_ISL_3187096, EPI_ISL_3187097, EPI_ISL_3187098, EPI_ISL_3187099, EPI_ISL_3187100, EPI_ISL_3187101, EPI_ISL_3187102, EPI_ISL_3187132, EPI_ISL_3187133, EPI_ISL_3187134, EPI_ISL_3187135, EPI_ISL_3187136, EPI_ISL_3187137, EPI_ISL_3187138, EPI_ISL_3187139, EPI_ISL_3187140, EPI_ISL_3187141, EPI_ISL_3187142, EPI_ISL_3187143, EPI_ISL_3187144, EPI_ISL_3187145, EPI_ISL_3187146, EPI_ISL_3187147, EPI_ISL_3187148, EPI_ISL_3187149, EPI_ISL_3187150, EPI_ISL_3187151, EPI_ISL_3187192, EPI_ISL_3187193, EPI_ISL_3187194, EPI_ISL_3187195, EPI_ISL_3187196, EPI_ISL_3187217, EPI_ISL_3281052, EPI_ISL_3281139, EPI_ISL_3281140, EPI_ISL_3281141, EPI_ISL_3281142, EPI_ISL_3281143, EPI_ISL_3281144, EPI_ISL_3281145, EPI_ISL_3281146, EPI_ISL_3281147, EPI_ISL_3281148, EPI_ISL_3281149, EPI_ISL_3281150, EPI_ISL_3281151, EPI_ISL_3281152, EPI_ISL_3281153, EPI_ISL_3281154, EPI_ISL_3281155, EPI_ISL_3281156, EPI_ISL_3281157, EPI_ISL_3281158, EPI_ISL_3281159, EPI_ISL_3281160, EPI_ISL_3281161, EPI_ISL_3281236, EPI_ISL_3281237, EPI_ISL_3281238, EPI_ISL_3281240, EPI_ISL_3281323, EPI_ISL_3281324, EPI_ISL_3281325, EPI_ISL_3281326, EPI_ISL_3281327, EPI_ISL_3281328, EPI_ISL_3281329, EPI_ISL_3281330, EPI_ISL_3281331, EPI_ISL_3281332, EPI_ISL_3281333, EPI_ISL_3281334, EPI_ISL_3281335, EPI_ISL_3281409, EPI_ISL_3281410, EPI_ISL_3281411, EPI_ISL_3305445, EPI_ISL_3305472, EPI_ISL_3305476, EPI_ISL_3305478, EPI_ISL_3305480, EPI_ISL_3305504, EPI_ISL_3305505, EPI_ISL_3398480, EPI_ISL_3398481, EPI_ISL_3398492, EPI_ISL_3398503, EPI_ISL_3398554, EPI_ISL_3398555, EPI_ISL_3398648, EPI_ISL_3398703, EPI_ISL_3398707, EPI_ISL_3398709, EPI_ISL_3398710, EPI_ISL_3398711, EPI_ISL_3398712, EPI_ISL_3398713, EPI_ISL_3401740, EPI_ISL_3401741, EPI_ISL_3401742, EPI_ISL_3401743, EPI_ISL_3401744, EPI_ISL_3401745, EPI_ISL_3401746, EPI_ISL_3401747, EPI_ISL_3401748, EPI_ISL_3401749, EPI_ISL_3401750 | | | |
| see above | Medlab Pathology | NSW Health Pathology - Institute of Clinical Pathology and Medical Research; Westmead Hospital; University of Sydney | Arnott A.; CIDM-PH et al.; Draper J.; Gall M.; Martinez E.; Rockett R.; Sintchenko V.; on behalf of ICPMR |
| EPI_ISL_779647, EPI_ISL_812434, EPI_ISL_854750, EPI_ISL_854754, EPI_ISL_854755, EPI_ISL_854756, EPI_ISL_854758, EPI_ISL_877570, EPI_ISL_877571, EPI_ISL_877572, EPI_ISL_877573, EPI_ISL_877574, EPI_ISL_877575, EPI_ISL_877580, EPI_ISL_877581, EPI_ISL_933778, EPI_ISL_962818, EPI_ISL_962819, EPI_ISL_962820, EPI_ISL_962821, EPI_ISL_962822, EPI_ISL_962827, EPI_ISL_979357, EPI_ISL_979358, EPI_ISL_979359, EPI_ISL_979360, EPI_ISL_979361, EPI_ISL_1033145, EPI_ISL_1033146, EPI_ISL_1033147, EPI_ISL_1033148, EPI_ISL_1033149, EPI_ISL_1033150, EPI_ISL_1033151, EPI_ISL_1033152, EPI_ISL_1033153, EPI_ISL_1033156, EPI_ISL_1033157, EPI_ISL_1061403, EPI_ISL_1249986, EPI_ISL_1249988, EPI_ISL_1249990, EPI_ISL_1249999, EPI_ISL_1250002, EPI_ISL_1250005, EPI_ISL_1913108, EPI_ISL_1913109, EPI_ISL_1913110, EPI_ISL_1913175, EPI_ISL_2321170, EPI_ISL_2321171, EPI_ISL_2321172, EPI_ISL_2321173, EPI_ISL_2321174, EPI_ISL_2321175, EPI_ISL_2321176, EPI_ISL_2321177, EPI_ISL_2321178, EPI_ISL_2321179, EPI_ISL_2321180, EPI_ISL_2321181, EPI_ISL_2379270, EPI_ISL_2379271, EPI_ISL_2379272, EPI_ISL_2379273, EPI_ISL_2379274, EPI_ISL_2379275, EPI_ISL_2379276, EPI_ISL_2379277, EPI_ISL_2379278, EPI_ISL_2379279, EPI_ISL_2379280, EPI_ISL_2379281, EPI_ISL_2379282, EPI_ISL_2379283, EPI_ISL_2379284, EPI_ISL_2379285, EPI_ISL_2379286, EPI_ISL_2379287, EPI_ISL_2379288, EPI_ISL_2379289, EPI_ISL_2379290, EPI_ISL_2379291, EPI_ISL_2379292, EPI_ISL_2405344, EPI_ISL_2405345, EPI_ISL_2405346, EPI_ISL_2405347, EPI_ISL_2405348, EPI_ISL_2405349, EPI_ISL_2405350, EPI_ISL_2405351, EPI_ISL_2405352, EPI_ISL_2405353, EPI_ISL_2405354, EPI_ISL_2405355, EPI_ISL_2405356, EPI_ISL_2405357, EPI_ISL_2405358, EPI_ISL_2405359, EPI_ISL_2482433, EPI_ISL_2482434, EPI_ISL_2482435, EPI_ISL_2482436, EPI_ISL_2482437, EPI_ISL_2482438, EPI_ISL_2482439, EPI_ISL_2482440, EPI_ISL_2482441, EPI_ISL_2482442, EPI_ISL_2482443, EPI_ISL_2482444, EPI_ISL_2482445, EPI_ISL_2482446, EPI_ISL_2482447, EPI_ISL_2482448, EPI_ISL_2482449, EPI_ISL_2482450, EPI_ISL_2482451, EPI_ISL_2482452, EPI_ISL_2482453, EPI_ISL_2482454, EPI_ISL_2482455, EPI_ISL_2482456, EPI_ISL_2482457, EPI_ISL_2482458, EPI_ISL_2482459, EPI_ISL_2482460, EPI_ISL_2482461, EPI_ISL_2598387, EPI_ISL_2598388, EPI_ISL_2598389, EPI_ISL_2598390, EPI_ISL_2598391, EPI_ISL_2598392, EPI_ISL_2598393, EPI_ISL_2598395, EPI_ISL_2598396, EPI_ISL_2598397, EPI_ISL_2598398, EPI_ISL_2598399, EPI_ISL_2598400, EPI_ISL_2598401, EPI_ISL_2688992, EPI_ISL_2688993, EPI_ISL_2688994, EPI_ISL_2688995, EPI_ISL_2839569, EPI_ISL_2839570, EPI_ISL_2839571, EPI_ISL_2839591, EPI_ISL_2839593, EPI_ISL_2907548, EPI_ISL_2907549, EPI_ISL_2907550, EPI_ISL_2907551, EPI_ISL_3030413, EPI_ISL_3030417, EPI_ISL_3030418, EPI_ISL_3030419, EPI_ISL_3030420, EPI_ISL_3030421, EPI_ISL_3030422, EPI_ISL_3030423, EPI_ISL_3030424, EPI_ISL_3030425, EPI_ISL_3050770, EPI_ISL_3050771, EPI_ISL_3050772, EPI_ISL_3050773, EPI_ISL_3050774, EPI_ISL_3050775, EPI_ISL_3050776, EPI_ISL_3050777, EPI_ISL_3050778, EPI_ISL_3050779, EPI_ISL_3050780, EPI_ISL_3050781, EPI_ISL_3050782, EPI_ISL_3050783, EPI_ISL_3050784, EPI_ISL_3050785, EPI_ISL_3050786, EPI_ISL_3050787, EPI_ISL_3050788, EPI_ISL_3050789, EPI_ISL_3050790, EPI_ISL_3050791, EPI_ISL_3050792, EPI_ISL_3050793, EPI_ISL_3050794, EPI_ISL_3050795, EPI_ISL_3050796, EPI_ISL_3050797, EPI_ISL_3050798, EPI_ISL_3050799, EPI_ISL_3050800, EPI_ISL_3050802, EPI_ISL_3050803, EPI_ISL_3050804, EPI_ISL_3050805, EPI_ISL_3050806, EPI_ISL_3050807, EPI_ISL_3050808, EPI_ISL_3050809, EPI_ISL_3050810, EPI_ISL_3050811, EPI_ISL_3050812, EPI_ISL_3050813, EPI_ISL_3134913, EPI_ISL_3134914, EPI_ISL_3134915, EPI_ISL_3134916, EPI_ISL_3134917 | | | |
| see above | Microbiological Diagnostic Unit - Public Health Laboratory (MDU-PHL) | MDU-PHL | Caly L.; Druce J.; M.L.; Meumann, E.; N.L.; Sait; Seemann T.; Sherry |
| EPI_ISL_2621569, EPI_ISL_2621572, EPI_ISL_2621573, EPI_ISL_2621574 | Microbiological Diagnostic Unit - Public Health Laboratory (MDU-PHL) | Microbiological Diagnostic Unit - Public Health Laboratory (MDU-PHL) | M.L.; N.L.; Sait; Seemann T.; Sherry |
| EPI_ISL_2250222, EPI_ISL_2250223, EPI_ISL_2250224, EPI_ISL_2250225, EPI_ISL_2250226, EPI_ISL_2250227, EPI_ISL_2250228, EPI_ISL_2250229, EPI_ISL_2250230, EPI_ISL_2250231, EPI_ISL_2250232, EPI_ISL_2250233 | | | |
| see above | Microbiological Diagnostic Unit - Public Health Laboratory (MDU-PHL) | Microbiological Diagnostic Unit Public Health Laboratory (MDU-PHL) | M.L.; N.L.; Sait; Seemann T.; Sherry |
| EPI_ISL_2659051 | Microbiological Diagnostic Unit - Public Health Laboratory (MDU-PHL) | Microbiological Diagnostics Unit Public Health Laboratory | M.L.; N.L.; Sait; Seemann T.; Sherry |
| EPI_ISL_1010699, EPI_ISL_1010700, EPI_ISL_1010701, EPI_ISL_1010702, EPI_ISL_1010703, EPI_ISL_1010704, EPI_ISL_1010707, EPI_ISL_1010708, EPI_ISL_1010709, EPI_ISL_1010710, EPI_ISL_1010711, EPI_ISL_1010712, EPI_ISL_1010713, EPI_ISL_1010714, EPI_ISL_1010715, EPI_ISL_1033024, EPI_ISL_1033025, EPI_ISL_1033026, EPI_ISL_1033027, EPI_ISL_1061033, EPI_ISL_1061034, EPI_ISL_1061035, EPI_ISL_1061036, EPI_ISL_1061037, EPI_ISL_1121977, EPI_ISL_1121978, EPI_ISL_1121979, EPI_ISL_1121981, EPI_ISL_1121982, EPI_ISL_1121983, EPI_ISL_1121984, EPI_ISL_1121985, EPI_ISL_1121986, EPI_ISL_1121987, EPI_ISL_1121988, EPI_ISL_1121989, EPI_ISL_1184094, EPI_ISL_1184095, EPI_ISL_1184096, EPI_ISL_1184097, EPI_ISL_1184098, EPI_ISL_1205303, EPI_ISL_1293045, EPI_ISL_1315066, EPI_ISL_1315067, EPI_ISL_1315068, EPI_ISL_1315069, EPI_ISL_1315070, EPI_ISL_1315071, EPI_ISL_1315072, EPI_ISL_1341506, EPI_ISL_1341507, EPI_ISL_1341508, EPI_ISL_1397953, EPI_ISL_1397954, EPI_ISL_1397955, EPI_ISL_1397956, EPI_ISL_1397957, EPI_ISL_1660398, EPI_ISL_1660399, EPI_ISL_1660400, EPI_ISL_1660401, EPI_ISL_1660402, EPI_ISL_1660403, EPI_ISL_1660404, EPI_ISL_1660405, EPI_ISL_1660406, EPI_ISL_1660407, EPI_ISL_1660408, EPI_ISL_1660409, EPI_ISL_1660410, EPI_ISL_1660411, EPI_ISL_1660476, EPI_ISL_1660477, EPI_ISL_1660478, EPI_ISL_1660479, EPI_ISL_1660480, EPI_ISL_1697278, EPI_ISL_1697279, EPI_ISL_1697280, EPI_ISL_1697281, EPI_ISL_1756021, EPI_ISL_1756022, EPI_ISL_1756023, EPI_ISL_1756024, EPI_ISL_1756025, EPI_ISL_1756026, EPI_ISL_1756027, EPI_ISL_1756028, EPI_ISL_1756029, EPI_ISL_1756030, EPI_ISL_1756031, EPI_ISL_1756032, EPI_ISL_1805698, EPI_ISL_1805699, EPI_ISL_1805700, EPI_ISL_2107442, EPI_ISL_2107443, EPI_ISL_2107444, EPI_ISL_2107445, EPI_ISL_2107446, EPI_ISL_2107447, EPI_ISL_2107448, EPI_ISL_2107449, EPI_ISL_2107450, EPI_ISL_2107451, EPI_ISL_2107452, EPI_ISL_2107453, EPI_ISL_2107454, EPI_ISL_2107455, EPI_ISL_2107456, EPI_ISL_2107457, EPI_ISL_2107458, EPI_ISL_2107459, EPI_ISL_2107499, EPI_ISL_2107500, EPI_ISL_2107501, EPI_ISL_2107502, EPI_ISL_2107503, EPI_ISL_2107504, EPI_ISL_2107505, EPI_ISL_2107506, EPI_ISL_2107529, EPI_ISL_2107530, EPI_ISL_2107531, EPI_ISL_2107532, EPI_ISL_2107533, EPI_ISL_2107534, EPI_ISL_2107535, EPI_ISL_2249256, EPI_ISL_2249257, EPI_ISL_2249258, EPI_ISL_2274250, EPI_ISL_2402459, EPI_ISL_2402460, EPI_ISL_2402461, EPI_ISL_2455225, EPI_ISL_2455226, EPI_ISL_2455227, EPI_ISL_2455228, EPI_ISL_2455229, EPI_ISL_2455230, EPI_ISL_2455231, EPI_ISL_2628996, EPI_ISL_2628997, EPI_ISL_2628998, EPI_ISL_2628999, EPI_ISL_2629000, EPI_ISL_2629001, EPI_ISL_2695797, EPI_ISL_2695798, EPI_ISL_2695799, EPI_ISL_2695800, EPI_ISL_2696273, EPI_ISL_2696274, EPI_ISL_2696275, EPI_ISL_2696276, EPI_ISL_2696277, EPI_ISL_2696278, EPI_ISL_2696279, EPI_ISL_2696280, EPI_ISL_2811796, EPI_ISL_2811797, EPI_ISL_2811798, EPI_ISL_2811799, EPI_ISL_2811800, EPI_ISL_2811801, EPI_ISL_2811802, EPI_ISL_2811803, EPI_ISL_2811804, EPI_ISL_2811806, EPI_ISL_2811807, EPI_ISL_2811808, EPI_ISL_2811809, EPI_ISL_2811810, EPI_ISL_2811811, EPI_ISL_2811812, EPI_ISL_2811813, EPI_ISL_2811814, EPI_ISL_2811815, EPI_ISL_2811816, EPI_ISL_2811817, EPI_ISL_2811818, EPI_ISL_2811819, EPI_ISL_2811820, EPI_ISL_2811821, EPI_ISL_2811822, EPI_ISL_2811823, EPI_ISL_2811824, EPI_ISL_2811825, EPI_ISL_2827952, EPI_ISL_2827953, EPI_ISL_2827954, EPI_ISL_2827955, EPI_ISL_2827956, EPI_ISL_2827957, EPI_ISL_2827958, EPI_ISL_2827959, EPI_ISL_2827960, EPI_ISL_2921613, EPI_ISL_2921614, EPI_ISL_2921615, EPI_ISL_2921616, EPI_ISL_2921617, EPI_ISL_2921618, EPI_ISL_2921619, EPI_ISL_2921620, EPI_ISL_2921621, EPI_ISL_2921622, EPI_ISL_2921843, EPI_ISL_2921844, EPI_ISL_2921845, EPI_ISL_3071989, EPI_ISL_3071990, EPI_ISL_3071991, EPI_ISL_3071992, EPI_ISL_3071993, EPI_ISL_3071994, EPI_ISL_3071995, EPI_ISL_3071996, EPI_ISL_3071997, EPI_ISL_3071998, EPI_ISL_3071999, EPI_ISL_3072000, EPI_ISL_3072001, EPI_ISL_3072002, EPI_ISL_3072003, EPI_ISL_3072004, EPI_ISL_3072005, EPI_ISL_3072006, EPI_ISL_3072007, EPI_ISL_3072008, EPI_ISL_3072009, EPI_ISL_3072010, EPI_ISL_3072011, EPI_ISL_3072012, EPI_ISL_3072013, EPI_ISL_3072014, EPI_ISL_3072015, EPI_ISL_3072016, EPI_ISL_3072017, EPI_ISL_3072018, EPI_ISL_3072019, EPI_ISL_3072020, EPI_ISL_3072021, EPI_ISL_3072022, EPI_ISL_3072023, EPI_ISL_3072024, EPI_ISL_3072025, EPI_ISL_3072026, EPI_ISL_3072027, EPI_ISL_3072028, EPI_ISL_3072029, EPI_ISL_3072030, EPI_ISL_3072031, EPI_ISL_3072032, EPI_ISL_3072033, EPI_ISL_3072034, EPI_ISL_3072036, EPI_ISL_3072037, EPI_ISL_3072038, EPI_ISL_3072039, EPI_ISL_3072040, EPI_ISL_3072041, EPI_ISL_3072042, EPI_ISL_3072043, EPI_ISL_3072044, EPI_ISL_3072045, EPI_ISL_3072046, EPI_ISL_3072047, EPI_ISL_3072048, EPI_ISL_3072049, EPI_ISL_3072050, EPI_ISL_3072051, EPI_ISL_3072052, EPI_ISL_3072053, EPI_ISL_3072054, EPI_ISL_3072056, EPI_ISL_3072057, EPI_ISL_3072058, EPI_ISL_3072059, EPI_ISL_3072060, EPI_ISL_3072062, EPI_ISL_3072063, EPI_ISL_3072064, EPI_ISL_3072065, EPI_ISL_3072066, EPI_ISL_3072067, EPI_ISL_3072068, EPI_ISL_3072069, EPI_ISL_3072070, EPI_ISL_3072071, EPI_ISL_3072073, EPI_ISL_3072074, EPI_ISL_3072075, EPI_ISL_3072076, EPI_ISL_3072077, EPI_ISL_3072078, EPI_ISL_3072079, EPI_ISL_3072080, EPI_ISL_3072081, EPI_ISL_3072082, EPI_ISL_3072083, EPI_ISL_3072084, EPI_ISL_3072085, EPI_ISL_3072086, EPI_ISL_3072088, EPI_ISL_3072089, EPI_ISL_3072090, EPI_ISL_3072091, EPI_ISL_3072092, EPI_ISL_3072093, EPI_ISL_3072095, EPI_ISL_3072096, EPI_ISL_3072097, EPI_ISL_3072099, EPI_ISL_3072100, EPI_ISL_3072102, EPI_ISL_3072103, EPI_ISL_3072106, EPI_ISL_3072107, EPI_ISL_3072108, EPI_ISL_3072110, EPI_ISL_3072112, EPI_ISL_3072113, EPI_ISL_3072115, EPI_ISL_3072116, EPI_ISL_3072117, EPI_ISL_3072118, EPI_ISL_3072119, EPI_ISL_3072120, EPI_ISL_3072121, EPI_ISL_3072123, EPI_ISL_3072124, EPI_ISL_3072125, EPI_ISL_3072126, EPI_ISL_3072127, EPI_ISL_3072128, EPI_ISL_3072129, EPI_ISL_3072130, EPI_ISL_3072131, EPI_ISL_3072132, EPI_ISL_3072133, EPI_ISL_3072134, EPI_ISL_3072135, EPI_ISL_3072136, EPI_ISL_3072137, EPI_ISL_3072138, EPI_ISL_3072140, EPI_ISL_3072141, EPI_ISL_3072142, EPI_ISL_3072145, EPI_ISL_3072146, EPI_ISL_3072148, EPI_ISL_3280812, EPI_ISL_3280813, EPI_ISL_3280814, EPI_ISL_3280815, EPI_ISL_3280816, EPI_ISL_3280817, EPI_ISL_3280818, EPI_ISL_3280819, EPI_ISL_3280820, EPI_ISL_3280821, EPI_ISL_3280822, EPI_ISL_3280823, EPI_ISL_3280824, EPI_ISL_3280825, EPI_ISL_3280826, EPI_ISL_3280827, EPI_ISL_3280828, EPI_ISL_3280829, EPI_ISL_3280830, EPI_ISL_3280831, EPI_ISL_3280832, EPI_ISL_3280833, EPI_ISL_3280834, EPI_ISL_3280835, EPI_ISL_3280836, EPI_ISL_3280837, EPI_ISL_3280838, EPI_ISL_3280839, EPI_ISL_3280927, EPI_ISL_3280928, EPI_ISL_3280929, EPI_ISL_3280930, EPI_ISL_3280931, EPI_ISL_3280932, EPI_ISL_3280933, EPI_ISL_3280934, EPI_ISL_3280935, EPI_ISL_3280936, EPI_ISL_3280937, EPI_ISL_3280938, EPI_ISL_3280939, EPI_ISL_3280940, EPI_ISL_3280941, EPI_ISL_3280942, EPI_ISL_3280943, EPI_ISL_3280944, EPI_ISL_3280945, EPI_ISL_3280946, EPI_ISL_3280947, EPI_ISL_3280948, EPI_ISL_3280949, EPI_ISL_3280950, EPI_ISL_3280951, EPI_ISL_3280952, EPI_ISL_3280953, EPI_ISL_3280954, EPI_ISL_3280955, EPI_ISL_3280956, EPI_ISL_3280957, EPI_ISL_3280958, EPI_ISL_3280959, EPI_ISL_3280960, EPI_ISL_3280961, EPI_ISL_3280962, EPI_ISL_3280963, EPI_ISL_3280964, EPI_ISL_3280965, EPI_ISL_3280966, EPI_ISL_3280967, EPI_ISL_3280968, EPI_ISL_3280969, EPI_ISL_3280970, EPI_ISL_3280971, EPI_ISL_3280972, EPI_ISL_3280973, EPI_ISL_3280974, EPI_ISL_3280975, EPI_ISL_3280976, EPI_ISL_3280977, EPI_ISL_3280978, EPI_ISL_3280979, EPI_ISL_3280980, EPI_ISL_3280981, EPI_ISL_3280982, EPI_ISL_3280983, EPI_ISL_3280984, EPI_ISL_3280985, EPI_ISL_3280986, EPI_ISL_3280987, EPI_ISL_3280988, EPI_ISL_3280989, EPI_ISL_3280990, EPI_ISL_3280991, EPI_ISL_3280992, EPI_ISL_3280993, EPI_ISL_3280994, EPI_ISL_3280995, EPI_ISL_3280996, EPI_ISL_3280997, EPI_ISL_3280998, EPI_ISL_3280999, EPI_ISL_3281000, EPI_ISL_3281001, EPI_ISL_3281002, EPI_ISL_3281003, EPI_ISL_3281004, EPI_ISL_3281005, EPI_ISL_3281006, EPI_ISL_3281007, EPI_ISL_3281008, EPI_ISL_3281009, EPI_ISL_3281010, EPI_ISL_3281011, EPI_ISL_3281012, EPI_ISL_3281013, EPI_ISL_3333179, EPI_ISL_3333180, EPI_ISL_3333181, EPI_ISL_3333182, EPI_ISL_3333183, EPI_ISL_3333184, EPI_ISL_3333185, EPI_ISL_3333186, EPI_ISL_3333187, EPI_ISL_3333188, EPI_ISL_3374155, EPI_ISL_3374156, EPI_ISL_3374157, EPI_ISL_3374158, EPI_ISL_3374159, EPI_ISL_3374160, EPI_ISL_3374161, EPI_ISL_3374162, EPI_ISL_3374163, EPI_ISL_3374164, EPI_ISL_3374165, EPI_ISL_3374166, EPI_ISL_3374167, EPI_ISL_3374168, EPI_ISL_3374169, EPI_ISL_3374170, EPI_ISL_3374171, EPI_ISL_3374172, EPI_ISL_3374173, EPI_ISL_3374174, EPI_ISL_3374175, EPI_ISL_3374176, EPI_ISL_3374177, EPI_ISL_3374178, EPI_ISL_3374179, EPI_ISL_3374180, EPI_ISL_3374181, EPI_ISL_3374182, EPI_ISL_3374183, EPI_ISL_3374184, EPI_ISL_3374185, EPI_ISL_3374186, EPI_ISL_3374187, EPI_ISL_3374188, EPI_ISL_3374189, EPI_ISL_3374190, EPI_ISL_3374191, EPI_ISL_3374192, EPI_ISL_3374193, EPI_ISL_3374194, EPI_ISL_3374195, EPI_ISL_3374196, EPI_ISL_3374197, EPI_ISL_3374198, EPI_ISL_3374199, EPI_ISL_3374200, EPI_ISL_3374201, EPI_ISL_3374202, EPI_ISL_3374203, EPI_ISL_3374204, EPI_ISL_3374205, EPI_ISL_3374206, EPI_ISL_3374207, EPI_ISL_3374208, EPI_ISL_3374209, EPI_ISL_3374210, EPI_ISL_3374211, EPI_ISL_3374212, EPI_ISL_3374213, EPI_ISL_3374214, EPI_ISL_3374215, EPI_ISL_3374216, EPI_ISL_3374217, EPI_ISL_3374218, EPI_ISL_3374219, EPI_ISL_3374220, EPI_ISL_3374221, EPI_ISL_3374222, EPI_ISL_3374223, EPI_ISL_3374224, EPI_ISL_3374225, EPI_ISL_3374226, EPI_ISL_3374227, EPI_ISL_3374228, EPI_ISL_3374230, EPI_ISL_3374231, EPI_ISL_3374232 | | | |
| see above | New South Wales Health Pathology Royal Prince Alfred Hospital | Microbiology RPAH | Au, J.; Bull, R.; Deveson, I.; Foster, C.; Rawlinson, W.; Ruiz Silva, M.; Van Hal, S. |
| EPI_ISL_968212, EPI_ISL_968213, EPI_ISL_1159377, EPI_ISL_1159378, EPI_ISL_1159379, EPI_ISL_1159380, EPI_ISL_1170949, EPI_ISL_1170950, EPI_ISL_1170951, EPI_ISL_1170952, EPI_ISL_1170953, EPI_ISL_1170954, EPI_ISL_1196413, EPI_ISL_1196414, EPI_ISL_1196415, EPI_ISL_1196416, EPI_ISL_1196417, EPI_ISL_1196418, EPI_ISL_1196419, EPI_ISL_1196420, EPI_ISL_1222761, EPI_ISL_1222762, EPI_ISL_1222763, EPI_ISL_1222764, EPI_ISL_1222765, EPI_ISL_1241835, EPI_ISL_1241836, EPI_ISL_1241837, EPI_ISL_1241838, EPI_ISL_1241839, EPI_ISL_1241840, EPI_ISL_1241841, EPI_ISL_1241842, EPI_ISL_1300523, EPI_ISL_1300524, EPI_ISL_1300534, EPI_ISL_1340868, EPI_ISL_1340869, EPI_ISL_1340870, EPI_ISL_1340871, EPI_ISL_1340872, EPI_ISL_1340874, EPI_ISL_1340875, EPI_ISL_1340876, EPI_ISL_1340877, EPI_ISL_1340878, EPI_ISL_1340879, EPI_ISL_1364538, EPI_ISL_1364539, EPI_ISL_1364541, EPI_ISL_1396519, EPI_ISL_1396520, EPI_ISL_1396521, EPI_ISL_1396524, EPI_ISL_1396525, EPI_ISL_1396526, EPI_ISL_1396527, EPI_ISL_1396530, EPI_ISL_1404914, EPI_ISL_1404920, EPI_ISL_1424004, EPI_ISL_1424005, EPI_ISL_1424006, EPI_ISL_1465880, EPI_ISL_1465881, EPI_ISL_1465882, EPI_ISL_1465883, EPI_ISL_1465885, EPI_ISL_1465886, EPI_ISL_1465889, EPI_ISL_1465890, EPI_ISL_1465891, EPI_ISL_1465893, EPI_ISL_1465894, EPI_ISL_1465895, EPI_ISL_1465896, EPI_ISL_1516885, EPI_ISL_1516886, EPI_ISL_1516887, EPI_ISL_1563660, EPI_ISL_1563665, EPI_ISL_1563670, EPI_ISL_1563675, EPI_ISL_1563678, EPI_ISL_1910856, EPI_ISL_1910857, EPI_ISL_1910859, EPI_ISL_1910860, EPI_ISL_2001058, EPI_ISL_2001059, EPI_ISL_2001060, EPI_ISL_2137032, EPI_ISL_2137033, EPI_ISL_2137034, EPI_ISL_2137035, EPI_ISL_2137036, EPI_ISL_2274244, EPI_ISL_2274245, EPI_ISL_2274246, EPI_ISL_2274247, EPI_ISL_2274248, EPI_ISL_2274249, EPI_ISL_2385968, EPI_ISL_2385969, EPI_ISL_2385970, EPI_ISL_2385971, EPI_ISL_2385972, EPI_ISL_2385973, EPI_ISL_2385974, EPI_ISL_2507084, EPI_ISL_2507085, EPI_ISL_2507086, EPI_ISL_2507087, EPI_ISL_2507088, EPI_ISL_2507089, EPI_ISL_2507090, EPI_ISL_2507091, EPI_ISL_2507092, EPI_ISL_2507093, EPI_ISL_2507094, EPI_ISL_2507095, EPI_ISL_2507096, EPI_ISL_2603817, EPI_ISL_2603818, EPI_ISL_2603819, EPI_ISL_2603820, EPI_ISL_2603821, EPI_ISL_2603822, EPI_ISL_2603823, EPI_ISL_2603824, EPI_ISL_2661514, EPI_ISL_2661515, EPI_ISL_2661516, EPI_ISL_2661517, EPI_ISL_2661518, EPI_ISL_2661519, EPI_ISL_2661520, EPI_ISL_2661521, EPI_ISL_2661522, EPI_ISL_2661523, EPI_ISL_2661524, EPI_ISL_2661525, EPI_ISL_2675252, EPI_ISL_2675253, EPI_ISL_2675254, EPI_ISL_2675255, EPI_ISL_2695786, EPI_ISL_2695787, EPI_ISL_2695788, EPI_ISL_2695789, EPI_ISL_2695790, EPI_ISL_2727636, EPI_ISL_2727637, EPI_ISL_2727638, EPI_ISL_2727639, EPI_ISL_2727640, EPI_ISL_2727641, EPI_ISL_2727642, EPI_ISL_2727643, EPI_ISL_2811889, EPI_ISL_2811890, EPI_ISL_2811891, EPI_ISL_2811892, EPI_ISL_2811893, EPI_ISL_2811894, EPI_ISL_2811895, EPI_ISL_2811896, EPI_ISL_2811897, EPI_ISL_2811898, EPI_ISL_2811899, EPI_ISL_2811900, EPI_ISL_2811901, EPI_ISL_2811902, EPI_ISL_2811903, EPI_ISL_2811904, EPI_ISL_2811905, EPI_ISL_2811906, EPI_ISL_2842837, EPI_ISL_2842838, EPI_ISL_2842839, EPI_ISL_2842840, EPI_ISL_2842841, EPI_ISL_2842842, EPI_ISL_2842843, EPI_ISL_2842844, EPI_ISL_2842845, EPI_ISL_2842846, EPI_ISL_2896242, EPI_ISL_2896243, EPI_ISL_2896244, EPI_ISL_2896245, EPI_ISL_2896246, EPI_ISL_2896247, EPI_ISL_2896248, EPI_ISL_2896249, EPI_ISL_2956008, EPI_ISL_2956009, EPI_ISL_2956010, EPI_ISL_2956011, EPI_ISL_2956012, EPI_ISL_2956013, EPI_ISL_2956014, EPI_ISL_3011177, EPI_ISL_3011178, EPI_ISL_3011179, EPI_ISL_3011180, EPI_ISL_3011181, EPI_ISL_3011182, EPI_ISL_3049190, EPI_ISL_3049191, EPI_ISL_3049192, EPI_ISL_3049193, EPI_ISL_3049194, EPI_ISL_3090751, EPI_ISL_3090752, EPI_ISL_3090753, EPI_ISL_3090754, EPI_ISL_3090755, EPI_ISL_3090756, EPI_ISL_3161812, EPI_ISL_3161813, EPI_ISL_3161814, EPI_ISL_3161815, EPI_ISL_3161816, EPI_ISL_3161817, EPI_ISL_3161818, EPI_ISL_3161819, EPI_ISL_3161820, EPI_ISL_3161821, EPI_ISL_3161822, EPI_ISL_3161823, EPI_ISL_3161824, EPI_ISL_3161825, EPI_ISL_3161826, EPI_ISL_3161827, EPI_ISL_3161828, EPI_ISL_3161829, EPI_ISL_3161830, EPI_ISL_3161831, EPI_ISL_3161832, EPI_ISL_3161833, EPI_ISL_3161834, EPI_ISL_3161835, EPI_ISL_3161836, EPI_ISL_3161837, EPI_ISL_3161838, EPI_ISL_3161839, EPI_ISL_3161840, EPI_ISL_3161841, EPI_ISL_3161842, EPI_ISL_3161843, EPI_ISL_3161844, EPI_ISL_3161845, EPI_ISL_3161846, EPI_ISL_3247178, EPI_ISL_3247179, EPI_ISL_3247180, EPI_ISL_3247181, EPI_ISL_3247182, EPI_ISL_3247183, EPI_ISL_3247184, EPI_ISL_3247185, EPI_ISL_3247186, EPI_ISL_3247187, EPI_ISL_3247188, EPI_ISL_3247189, EPI_ISL_3247190, EPI_ISL_3247191, EPI_ISL_3247192, EPI_ISL_3247193, EPI_ISL_3247194, EPI_ISL_3247195, EPI_ISL_3247196, EPI_ISL_3247197, EPI_ISL_3247198, EPI_ISL_3247199, EPI_ISL_3247200, EPI_ISL_3247201, EPI_ISL_3247202, EPI_ISL_3247203, EPI_ISL_3247204, EPI_ISL_3247205, EPI_ISL_3247206, EPI_ISL_3247207, EPI_ISL_3247208, EPI_ISL_3247209, EPI_ISL_3247210, EPI_ISL_3247211, EPI_ISL_3247212, EPI_ISL_3247213, EPI_ISL_3247214, EPI_ISL_3247215, EPI_ISL_3247216, EPI_ISL_3247217, EPI_ISL_3247218, EPI_ISL_3247219, EPI_ISL_3247220, EPI_ISL_3247221, EPI_ISL_3247222, EPI_ISL_3247223, EPI_ISL_3247224, EPI_ISL_3247225, EPI_ISL_3247226, EPI_ISL_3247227, EPI_ISL_3247228, EPI_ISL_3247229, EPI_ISL_3247230, EPI_ISL_3247231, EPI_ISL_3247232, EPI_ISL_3247233, EPI_ISL_3333190, EPI_ISL_3333191, EPI_ISL_3333192, EPI_ISL_3333193, EPI_ISL_3333194, EPI_ISL_3333195, EPI_ISL_3333196, EPI_ISL_3333197, EPI_ISL_3333198, EPI_ISL_3333199, EPI_ISL_3333200, EPI_ISL_3333201, EPI_ISL_3333202, EPI_ISL_3333203, EPI_ISL_3333204, EPI_ISL_3333206, EPI_ISL_3333207, EPI_ISL_3333208, EPI_ISL_3333209, EPI_ISL_3333210, EPI_ISL_3333211, EPI_ISL_3333212, EPI_ISL_3333213, EPI_ISL_3333215, EPI_ISL_3333216, EPI_ISL_3333217, EPI_ISL_3333218, EPI_ISL_3333219, EPI_ISL_3333220, EPI_ISL_3333221, EPI_ISL_3333222, EPI_ISL_3333223, EPI_ISL_3333224, EPI_ISL_3333225, EPI_ISL_3333226, EPI_ISL_3333227, EPI_ISL_3333229, EPI_ISL_3333230, EPI_ISL_3333231 | | | |
| see above | PHV-FSS | PHV-FSS | Chenwei Wang on behalf of Q-PHIRE Genomics; Chenwei-Wang; Q-PHIRE Genomics; Son Nguyen; Son Nguyen et al; Son Nguyen et al. |
| EPI_ISL_2662451, EPI_ISL_2713075, EPI_ISL_2758809, EPI_ISL_2932263 | PathWest Laboratory Medicine WA | PathWest Laboratory Medicine WA | PathWest Laboratory Medicine WA; PathWest Laboratory Medicine WA Microbial Surveillance Unit |
| EPI_ISL_1911192 | PathWest Laboratory Medicine WA | PathWest Laboratory Medicine WA Microbial Surveillance UNit | PathWest Laboratory Medicine WA Microbial Surveillance UNit |
| EPI_ISL_794675, EPI_ISL_794676, EPI_ISL_810964, EPI_ISL_810966, EPI_ISL_810967, EPI_ISL_810968, EPI_ISL_810969, EPI_ISL_857466, EPI_ISL_857467, EPI_ISL_857468, EPI_ISL_857486, EPI_ISL_857487, EPI_ISL_857488, EPI_ISL_933787, EPI_ISL_933788, EPI_ISL_933790, EPI_ISL_933791, EPI_ISL_933792, EPI_ISL_933793, EPI_ISL_933794, EPI_ISL_933795, EPI_ISL_933796, EPI_ISL_933798, EPI_ISL_933799, EPI_ISL_933800, EPI_ISL_933801, EPI_ISL_933802, EPI_ISL_1017678, EPI_ISL_1069390, EPI_ISL_1069391, EPI_ISL_1069392, EPI_ISL_1069393, EPI_ISL_1184507, EPI_ISL_1198800, EPI_ISL_1198801, EPI_ISL_1198802, EPI_ISL_1198803, EPI_ISL_1295930, EPI_ISL_1295931, EPI_ISL_1295932, EPI_ISL_1295933, EPI_ISL_1295934, EPI_ISL_1295935, EPI_ISL_1295936, EPI_ISL_1295937, EPI_ISL_1366739, EPI_ISL_1366740, EPI_ISL_1384811, EPI_ISL_1384812, EPI_ISL_1416321, EPI_ISL_1416322, EPI_ISL_1416323, EPI_ISL_1416324, EPI_ISL_1416325, EPI_ISL_1416326, EPI_ISL_1448426, EPI_ISL_1448427, EPI_ISL_1448428, EPI_ISL_1448429, EPI_ISL_1448430, EPI_ISL_1508992, EPI_ISL_1508993, EPI_ISL_1508994, EPI_ISL_1508995, EPI_ISL_1508996, EPI_ISL_1508997, EPI_ISL_1620802, EPI_ISL_1620803, | | | |

| | | | |
|---|---|---|---|
| EPI_ISL_1620804, EPI_ISL_1620805, EPI_ISL_1620806, EPI_ISL_1620807, EPI_ISL_1633674, EPI_ISL_1633675, EPI_ISL_1633676, EPI_ISL_1633677, EPI_ISL_1633678, EPI_ISL_1633679, EPI_ISL_1633680, EPI_ISL_1633681, EPI_ISL_1633682, EPI_ISL_1633683, EPI_ISL_1669116, EPI_ISL_1669117, EPI_ISL_1669118, EPI_ISL_1669119, EPI_ISL_1669120, EPI_ISL_1669121, EPI_ISL_1669122, EPI_ISL_1669123, EPI_ISL_1669124, EPI_ISL_1669125, EPI_ISL_1669126, EPI_ISL_1750963, EPI_ISL_1750964, EPI_ISL_1750965, EPI_ISL_1750966, EPI_ISL_1750967, EPI_ISL_1750968, EPI_ISL_1816917, EPI_ISL_1816918, EPI_ISL_1816919, EPI_ISL_1816920, EPI_ISL_1816921, EPI_ISL_1816922, EPI_ISL_1816923, EPI_ISL_1816924, EPI_ISL_1817558, EPI_ISL_1817559, EPI_ISL_1817560, EPI_ISL_1828698, EPI_ISL_1828699, EPI_ISL_1828700, EPI_ISL_1914666, EPI_ISL_1914667, EPI_ISL_1914668, EPI_ISL_1914669, EPI_ISL_1972908, EPI_ISL_1972909, EPI_ISL_1972910, EPI_ISL_1972911, EPI_ISL_1972912, EPI_ISL_2081529, EPI_ISL_2098723, EPI_ISL_2226229, EPI_ISL_2304306, EPI_ISL_2322898, EPI_ISL_2322899, EPI_ISL_2598296, EPI_ISL_2598297, EPI_ISL_2757655, EPI_ISL_2757656, EPI_ISL_2757657, EPI_ISL_2784344, EPI_ISL_2841828, EPI_ISL_2841829, EPI_ISL_2841830, EPI_ISL_2841831, EPI_ISL_2861270, EPI_ISL_2942965, EPI_ISL_2942966, EPI_ISL_2942967, EPI_ISL_2982303, EPI_ISL_3038898, EPI_ISL_3038899, EPI_ISL_3150198, EPI_ISL_3150199, EPI_ISL_3150200, EPI_ISL_3150201, EPI_ISL_3220302, EPI_ISL_3245741, EPI_ISL_3245742, EPI_ISL_3245743, EPI_ISL_3245744, EPI_ISL_3245745, EPI_ISL_3245746, EPI_ISL_3245747, EPI_ISL_3245748, EPI_ISL_3245749, EPI_ISL_3245750, EPI_ISL_3245751, EPI_ISL_3245752, EPI_ISL_3245753, EPI_ISL_3245754 | | | |
| see above | PathWest Laboratory Medicine WA | PathWest Laboratory Medicine WA Microbial Surveillance Unit | PathWest Laboratory Medicine WA Microbial Surveillance Unit |
| EPI_ISL_2691707 | PathWest Laboratory Medicine WA Microbial Surveillance Unit | PathWest Laboratory Medicine WA Microbial Surveillance Unit | Hospital Ave; Nedlands WA 6009; QEII Medical Centre |
| EPI_ISL_3281036 | Pathology North - Gosford Hospital - NSW Health Pathology | NSW Health Pathology - Institute of Clinical Pathology and Medical Research; Westmead Hospital; University of Sydney | Arnott A.; Draper J.; Gall M.; Martinez E.; Rockett R.; Sintchenko V.; on behalf of ICPMR |
| EPI_ISL_2721376, EPI_ISL_3401685, EPI_ISL_3401784, EPI_ISL_3401785, EPI_ISL_3401787, EPI_ISL_3401788, EPI_ISL_3401789, EPI_ISL_3401842, EPI_ISL_3401843, EPI_ISL_3401844, EPI_ISL_3401845, EPI_ISL_3401846, EPI_ISL_3401847 | | | |
| see above | Pathology North - Hunter - NSW Health Pathology | NSW Health Pathology - Institute of Clinical Pathology and Medical Research; Westmead Hospital; University of Sydney | Arnott A.; CIDM-PH et al.; Draper J.; Gall M.; Martinez E.; Rockett R.; Sintchenko V.; on behalf of ICPMR |
| EPI_ISL_1904445 | Pathology North - Lismore laboratory - NSW Health Pathology | NSW Health Pathology - Institute of Clinical Pathology and Medical Research; Westmead Hospital; University of Sydney | CIDM-PH et al. |
| EPI_ISL_812521, EPI_ISL_2812817, EPI_ISL_3186765, EPI_ISL_3186910, EPI_ISL_3186911, EPI_ISL_3186912, EPI_ISL_3186913, EPI_ISL_3187159, EPI_ISL_3281043, EPI_ISL_3281059, EPI_ISL_3281241, EPI_ISL_3281336, EPI_ISL_3281337, EPI_ISL_3398589, EPI_ISL_3398714, EPI_ISL_3401848 | | | |
| see above | Pathology North - Royal North Shore Hospital - NSW Health Pathology | NSW Health Pathology - Institute of Clinical Pathology and Medical Research; Westmead Hospital; University of Sydney | Arnott A.; CIDM-PH et al.; Draper J.; Gall M.; Martinez E.; Rockett R.; Sintchenko V.; on behalf of ICPMR |
| EPI_ISL_767937, EPI_ISL_803108, EPI_ISL_803109, EPI_ISL_2812823, EPI_ISL_2812846, EPI_ISL_2812892, EPI_ISL_2812893, EPI_ISL_2812894, EPI_ISL_2812895, EPI_ISL_2812902, EPI_ISL_2828058, EPI_ISL_2894148, EPI_ISL_2894151, EPI_ISL_3007297, EPI_ISL_3015124, EPI_ISL_3015127, EPI_ISL_3015147, EPI_ISL_3184891, EPI_ISL_3184928, EPI_ISL_3186217, EPI_ISL_3186259, EPI_ISL_3186260, EPI_ISL_3186261, EPI_ISL_3186291, EPI_ISL_3186639, EPI_ISL_3186719, EPI_ISL_3186951, EPI_ISL_3187017, EPI_ISL_3187018, EPI_ISL_3187019, EPI_ISL_3187166, EPI_ISL_3281053, EPI_ISL_3281232, EPI_ISL_3281396, EPI_ISL_3281397, EPI_ISL_3281398, EPI_ISL_3281399, EPI_ISL_3281400, EPI_ISL_3281401, EPI_ISL_3281412, EPI_ISL_3305407, EPI_ISL_3398404, EPI_ISL_3398439, EPI_ISL_3398482, EPI_ISL_3398519, EPI_ISL_3398520, EPI_ISL_3401849 | | | |
| see above | Pathology West - NSW Health Pathology | NSW Health Pathology - Institute of Clinical Pathology and Medical Research; Westmead Hospital; University of Sydney | Arnott A.; CIDM-PH et al.; Draper J.; Gall M.; Martinez E.; Rockett R.; Sintchenko V.; on behalf of ICPMR |
| EPI_ISL_849750, EPI_ISL_849751, EPI_ISL_849752, EPI_ISL_849753, EPI_ISL_849754, EPI_ISL_849755, EPI_ISL_849756, EPI_ISL_849757, EPI_ISL_849758, EPI_ISL_849759, EPI_ISL_849760, EPI_ISL_849761, EPI_ISL_849762 | | | |
| see above | Public Health Virology Laboratory, Forensic and Scientific Services (PHV-FSS) | Public Health Virology Laboratory, Forensic and Scientific Services (PHV-FSS) | Son Nguyen et al |
| EPI_ISL_985234, EPI_ISL_985235, EPI_ISL_1061309, EPI_ISL_1061310, EPI_ISL_1098644, EPI_ISL_1098645, EPI_ISL_1121039, EPI_ISL_1121040, EPI_ISL_1137503, EPI_ISL_1137504, EPI_ISL_1137505, EPI_ISL_1804614, EPI_ISL_1910858, EPI_ISL_2433926, EPI_ISL_2433928 | | | |
| see above | Public Health Virology-Forensic and Scientific Services | Public Health Virology-Forensic and Scientific Services | Alyssa T. Pyke; Alyssa T. Pyke et al.; Son Nguyen; Son Nguyen et al |
| EPI_ISL_944644, EPI_ISL_944738, EPI_ISL_944739, EPI_ISL_944740, EPI_ISL_944741, EPI_ISL_944742, EPI_ISL_944743, EPI_ISL_944744, EPI_ISL_944745, EPI_ISL_944746, EPI_ISL_944747, EPI_ISL_944748, EPI_ISL_962180, EPI_ISL_1273076, EPI_ISL_1273077, EPI_ISL_1273078, EPI_ISL_1273079, EPI_ISL_1273080, EPI_ISL_1273081, EPI_ISL_1273082, EPI_ISL_1273083, EPI_ISL_1300526, EPI_ISL_1300529, EPI_ISL_1300532, EPI_ISL_1300533, EPI_ISL_1306121, EPI_ISL_1306122, EPI_ISL_1306123, EPI_ISL_1306124, EPI_ISL_1306125, EPI_ISL_1306126, EPI_ISL_1306127, EPI_ISL_1306128, EPI_ISL_1306129, EPI_ISL_1306130, EPI_ISL_1306131, EPI_ISL_1306132, EPI_ISL_1306134, EPI_ISL_1306135, EPI_ISL_1306136, EPI_ISL_1306137, EPI_ISL_1306138, EPI_ISL_1340873, EPI_ISL_1364540, EPI_ISL_1396522, EPI_ISL_1396523, EPI_ISL_1396528, EPI_ISL_1396529, EPI_ISL_1396531, EPI_ISL_1396532, EPI_ISL_1396533, EPI_ISL_1404915, EPI_ISL_1404916, EPI_ISL_1404917, EPI_ISL_1404918, EPI_ISL_1404919, EPI_ISL_1404921, EPI_ISL_1404922, EPI_ISL_1424002, EPI_ISL_1424003, EPI_ISL_1465887, EPI_ISL_1465888, EPI_ISL_1465892, EPI_ISL_1483027, EPI_ISL_1483028, EPI_ISL_1483029, EPI_ISL_1483030, EPI_ISL_1483031, EPI_ISL_1516882, EPI_ISL_1516883, EPI_ISL_1516884, EPI_ISL_1516888, EPI_ISL_1516889, EPI_ISL_1633656, EPI_ISL_1633657, EPI_ISL_1633658, EPI_ISL_1633659, EPI_ISL_1754856, EPI_ISL_1754857, EPI_ISL_1754858, EPI_ISL_1754859, EPI_ISL_1754860, EPI_ISL_1754861, EPI_ISL_1754862, EPI_ISL_1754863, EPI_ISL_1754864, EPI_ISL_1754865, EPI_ISL_1754866, EPI_ISL_1754867, EPI_ISL_1803794 | | | |
| see above | Public Health Virology-Forensic and Scientific Services (PHV-FSS) | Public Health Virology-Forensic and Scientific Services (PHV-FSS) | Alyssa T. Pyke et al.; Son Nguyen; Son Nguyen et al |
| EPI_ISL_779409, EPI_ISL_779410, EPI_ISL_779411, EPI_ISL_812423, EPI_ISL_854745, EPI_ISL_2321157, EPI_ISL_2321158, EPI_ISL_2321159, EPI_ISL_2321160, EPI_ISL_2321161, EPI_ISL_2321162, EPI_ISL_2321163, EPI_ISL_2321164, EPI_ISL_2321165, EPI_ISL_2321166, EPI_ISL_2321167, EPI_ISL_2321168, EPI_ISL_2321169, EPI_ISL_2379269, EPI_ISL_2688991, EPI_ISL_2839562, EPI_ISL_2839563, EPI_ISL_2839567, EPI_ISL_2839568, EPI_ISL_2907544, EPI_ISL_2907545, EPI_ISL_2907546, EPI_ISL_3030412, EPI_ISL_3134918 | | | |
| see above | Royal Darwin Hospital Pathology | MDU-PHL | Caly L.; Druce J.; M.L.; Meumann, E.; N.L.; Sait; Seemann T.; Sherry |
| EPI_ISL_2250185, EPI_ISL_2250186, EPI_ISL_2250187, EPI_ISL_2250188, EPI_ISL_2250189, EPI_ISL_2250190, EPI_ISL_2250191, EPI_ISL_2250192, EPI_ISL_2250193, EPI_ISL_2250194, EPI_ISL_2250195, EPI_ISL_2250196, EPI_ISL_2250197, EPI_ISL_2250198, EPI_ISL_2250199, EPI_ISL_2250200, EPI_ISL_2250201, EPI_ISL_2250202, EPI_ISL_2250203, EPI_ISL_2250204, EPI_ISL_2250205, EPI_ISL_2250206, EPI_ISL_2250207, EPI_ISL_2250208, EPI_ISL_2250209, EPI_ISL_2250210, EPI_ISL_2250211, EPI_ISL_2250212, EPI_ISL_2250213, EPI_ISL_2250214, EPI_ISL_2250215, EPI_ISL_2250216, EPI_ISL_2250217, EPI_ISL_2250218, EPI_ISL_2250219, EPI_ISL_2250220 | | | |
| see above | Royal Darwin Hospital Pathology | Microbiological Diagnostic Unit Public Health Laboratory (MDU-PHL) | Caly L.; Druce J.; M.L.; Meumann, E.; N.L.; Sait; Seemann T.; Sherry |
| EPI_ISL_3247060 | Royal Hobart Hospital | Royal Hobart Hospital | Dr L. Cooley; Mr R Vanhaeften |
| EPI_ISL_771368, EPI_ISL_771369, EPI_ISL_771370, EPI_ISL_812356, EPI_ISL_812357, EPI_ISL_812358, EPI_ISL_855344, EPI_ISL_1029956, EPI_ISL_1229178, EPI_ISL_1701832, EPI_ISL_1704673, EPI_ISL_1704674, EPI_ISL_1704675, EPI_ISL_1704676, EPI_ISL_1704677, EPI_ISL_1704678, EPI_ISL_1704786, EPI_ISL_1704787, EPI_ISL_1704788, EPI_ISL_1704789, EPI_ISL_1704790, EPI_ISL_1704791, EPI_ISL_1704792, EPI_ISL_1704793, EPI_ISL_1704794, EPI_ISL_1704795, EPI_ISL_1704796, EPI_ISL_1704797, EPI_ISL_1704798, EPI_ISL_1704799, EPI_ISL_1704800, EPI_ISL_1704801, EPI_ISL_1704802, EPI_ISL_1704803, EPI_ISL_1704804, EPI_ISL_1704805, EPI_ISL_1704806, EPI_ISL_1704807, EPI_ISL_1704808, EPI_ISL_1706364, EPI_ISL_1706365, EPI_ISL_1706366, EPI_ISL_1706367, EPI_ISL_2462337, EPI_ISL_2462338, EPI_ISL_2462339, EPI_ISL_2462340, EPI_ISL_2462341, EPI_ISL_2462342, EPI_ISL_2462343, EPI_ISL_2462344, EPI_ISL_2462345, EPI_ISL_2462346, EPI_ISL_2462347, EPI_ISL_2462348, EPI_ISL_2462349, EPI_ISL_2462350, EPI_ISL_2462351, EPI_ISL_2462352, EPI_ISL_2462353, EPI_ISL_2462354, EPI_ISL_2462355, EPI_ISL_2462356, EPI_ISL_2462357, EPI_ISL_2462358, EPI_ISL_2462359, EPI_ISL_2462360, EPI_ISL_2462361, EPI_ISL_2462362, EPI_ISL_2462363, EPI_ISL_2462364, EPI_ISL_2462365, EPI_ISL_2462366, EPI_ISL_2462367, EPI_ISL_2462368, EPI_ISL_2839979, EPI_ISL_2839980, EPI_ISL_2839981, EPI_ISL_2839982, EPI_ISL_2839983, EPI_ISL_2839984, EPI_ISL_2839985, EPI_ISL_2839986, EPI_ISL_2839987, EPI_ISL_2839988, EPI_ISL_2839989, EPI_ISL_2839990, EPI_ISL_2839991, EPI_ISL_2839992, EPI_ISL_2839993, EPI_ISL_2839994, EPI_ISL_2839995, EPI_ISL_2839996, EPI_ISL_2839997, EPI_ISL_2839998, EPI_ISL_2978690, EPI_ISL_2978692, EPI_ISL_2978694, EPI_ISL_2978696, EPI_ISL_2978698, EPI_ISL_2978700, EPI_ISL_3071861, EPI_ISL_3071862, EPI_ISL_3071863, EPI_ISL_3071864, EPI_ISL_3071865, EPI_ISL_3071866, EPI_ISL_3391988, EPI_ISL_3391989, EPI_ISL_3391990, EPI_ISL_3391991, EPI_ISL_3391992 | | | |
| see above | SA Pathology | SA Pathology | Caitlin Selway; Chuan Kok Lim; Geoff Higgins; Ivan Bastian; Julien Soubrier; Karin Kassahn; Lex Leong; Mark Turra; Song Gao |
| EPI_ISL_779401, EPI_ISL_779402, EPI_ISL_791279, EPI_ISL_792037, EPI_ISL_845797, EPI_ISL_857308, EPI_ISL_857310, EPI_ISL_857311, EPI_ISL_857312, EPI_ISL_857313, EPI_ISL_872575, EPI_ISL_872577, EPI_ISL_872582, EPI_ISL_872583, EPI_ISL_872584, EPI_ISL_942895, EPI_ISL_1098648, EPI_ISL_1098649, EPI_ISL_1098650, EPI_ISL_1098651, EPI_ISL_1098652, EPI_ISL_1383236, EPI_ISL_1383237, EPI_ISL_1383238, EPI_ISL_1383239, EPI_ISL_1383240, EPI_ISL_1383241, EPI_ISL_1383242, EPI_ISL_1383243, EPI_ISL_1383244, EPI_ISL_1383245, EPI_ISL_1383247, EPI_ISL_1383248, EPI_ISL_1447288, EPI_ISL_1447289, EPI_ISL_1447290, EPI_ISL_1447291, EPI_ISL_1904449, EPI_ISL_1904450, EPI_ISL_1904451, EPI_ISL_1904452, EPI_ISL_1904453, EPI_ISL_1904454, EPI_ISL_1904455, EPI_ISL_1904456, EPI_ISL_1904457, EPI_ISL_1904458, EPI_ISL_1904459, EPI_ISL_1904460, EPI_ISL_1904461, EPI_ISL_1904462, EPI_ISL_1904463, EPI_ISL_1904464, EPI_ISL_1904465, EPI_ISL_1904466, EPI_ISL_1904468, EPI_ISL_1904469, EPI_ISL_1904470, EPI_ISL_1904471, EPI_ISL_2405109, EPI_ISL_2405110, EPI_ISL_2405113, EPI_ISL_2405114, EPI_ISL_2405115, EPI_ISL_2405116, EPI_ISL_2405117, EPI_ISL_2462407, EPI_ISL_2462408, EPI_ISL_2462410, EPI_ISL_2462412, EPI_ISL_2462413, EPI_ISL_2462414, EPI_ISL_2462416, EPI_ISL_2462418, EPI_ISL_2462419, EPI_ISL_2462420, EPI_ISL_2462422, EPI_ISL_2462423, EPI_ISL_2462424, EPI_ISL_2462425, EPI_ISL_2462430, EPI_ISL_2462431, EPI_ISL_2462432, EPI_ISL_2462433, EPI_ISL_2462434, EPI_ISL_2650451, EPI_ISL_2650452, EPI_ISL_2650453, EPI_ISL_2650454, EPI_ISL_2650455, EPI_ISL_2650456, EPI_ISL_2650457, EPI_ISL_2650458, EPI_ISL_2650459, EPI_ISL_2650460, EPI_ISL_2650461, EPI_ISL_2650462, EPI_ISL_2650463, EPI_ISL_2650464, EPI_ISL_2650465, EPI_ISL_2650466, EPI_ISL_2828016, EPI_ISL_2828017, EPI_ISL_2828018, EPI_ISL_2828069, EPI_ISL_2828070, EPI_ISL_2828071, EPI_ISL_2828072, EPI_ISL_2828073, EPI_ISL_2828074, EPI_ISL_3305409 | | | |
| see above | South Eastern Area Laboratory Services (SEALS) | NSW Health Pathology - Institute of Clinical Pathology and Medical Research; Westmead Hospital; University of Sydney | Arnott A.; CIDM-PH et al.; Draper J.; Gall M.; Martinez E.; Rockett R.; Sintchenko V.; on behalf of ICPMR |
| EPI_ISL_2861251, EPI_ISL_2861252 | Southern IML | NSW Health Pathology - Institute of Clinical Pathology and Medical Research; Westmead Hospital; University of Sydney | CIDM-PH et al. |
| EPI_ISL_2812814 | Southern IML Pathology | NSW Health Pathology - Institute of Clinical Pathology and Medical Research; Westmead Hospital; University of Sydney | CIDM-PH et al. |
| EPI_ISL_2812826, EPI_ISL_2894137, EPI_ISL_3185074, EPI_ISL_3187103, EPI_ISL_3398715, EPI_ISL_3398716 | Southern. IML Pathology | NSW Health Pathology - Institute of Clinical Pathology and Medical Research; Westmead Hospital; University of Sydney | Arnott A.; CIDM-PH et al.; Draper J.; Gall M.; Martinez E.; Rockett R.; Sintchenko V.; on behalf of ICPMR |
| EPI_ISL_872581, EPI_ISL_2680982, EPI_ISL_2680983, EPI_ISL_2754121, EPI_ISL_2786933, EPI_ISL_2786934, EPI_ISL_2811985, EPI_ISL_2811996, EPI_ISL_2812819, EPI_ISL_2812842, EPI_ISL_2812843, EPI_ISL_2812844, EPI_ISL_2812845, EPI_ISL_2812859, EPI_ISL_2812860, EPI_ISL_2812861, EPI_ISL_2812862, EPI_ISL_2812863, EPI_ISL_2812864, EPI_ISL_2812865, EPI_ISL_2812866, EPI_ISL_2812867, EPI_ISL_2812868, EPI_ISL_2812869, EPI_ISL_2812878, EPI_ISL_2812879, EPI_ISL_2812885, EPI_ISL_2812891, EPI_ISL_2812918, EPI_ISL_2812919, EPI_ISL_2861255, EPI_ISL_2876015, EPI_ISL_2876016, EPI_ISL_2876017, EPI_ISL_2876018, EPI_ISL_2876019, EPI_ISL_2876020, EPI_ISL_2876021, EPI_ISL_2876022, EPI_ISL_2876023, EPI_ISL_2876024, EPI_ISL_2876025, EPI_ISL_2876038, EPI_ISL_2876039, EPI_ISL_2876040, EPI_ISL_2876041, EPI_ISL_2876042, EPI_ISL_2877038, EPI_ISL_3007295, EPI_ISL_3015130, EPI_ISL_3015131, EPI_ISL_3015132, EPI_ISL_3015133, EPI_ISL_3015134, EPI_ISL_3015135, EPI_ISL_3015136, EPI_ISL_3015137, EPI_ISL_3015138, EPI_ISL_3015139, EPI_ISL_3015140, EPI_ISL_3015163, EPI_ISL_3015164, EPI_ISL_3015165, EPI_ISL_3015166, EPI_ISL_3015167, EPI_ISL_3015168, EPI_ISL_3015169, EPI_ISL_3119465, EPI_ISL_3184886, EPI_ISL_3184901, EPI_ISL_3184902, EPI_ISL_3184915, EPI_ISL_3184916, EPI_ISL_3184918, EPI_ISL_3184926, EPI_ISL_3184931, EPI_ISL_3184934, EPI_ISL_3184947, EPI_ISL_3184948, EPI_ISL_3184949, EPI_ISL_3184950, EPI_ISL_3184952, EPI_ISL_3184957, EPI_ISL_3184958, EPI_ISL_3184959, EPI_ISL_3184960, EPI_ISL_3184961, EPI_ISL_3184962, EPI_ISL_3184963, EPI_ISL_3184964, EPI_ISL_3184965, EPI_ISL_3184966, EPI_ISL_3184967, EPI_ISL_3184968, EPI_ISL_3184969, EPI_ISL_3184970, EPI_ISL_3184971, EPI_ISL_3185047, EPI_ISL_3185048, EPI_ISL_3185075, EPI_ISL_3185076, EPI_ISL_3185077, EPI_ISL_3185078, EPI_ISL_3185079, EPI_ISL_3185080, EPI_ISL_3185081, EPI_ISL_3185082, EPI_ISL_3185083, EPI_ISL_3185084, EPI_ISL_3185085, EPI_ISL_3185086, EPI_ISL_3185087, EPI_ISL_3185088, EPI_ISL_3185089, EPI_ISL_3185090, EPI_ISL_3185091, EPI_ISL_3185092, EPI_ISL_3185093, EPI_ISL_3185094, EPI_ISL_3185095, EPI_ISL_3185096, EPI_ISL_3185097, EPI_ISL_3185098, EPI_ISL_3185099, EPI_ISL_3185100, EPI_ISL_3185101, EPI_ISL_3185102, EPI_ISL_3185117, EPI_ISL_3185118, EPI_ISL_3185119, EPI_ISL_3185120, EPI_ISL_3185121, EPI_ISL_3185122, EPI_ISL_3185123, EPI_ISL_3185124, EPI_ISL_3185125, EPI_ISL_3185126, EPI_ISL_3185127, EPI_ISL_3185128, EPI_ISL_3185129, EPI_ISL_3185130, EPI_ISL_3185131, EPI_ISL_3185132, EPI_ISL_3185133, EPI_ISL_3185134, EPI_ISL_3185135, EPI_ISL_3185193, EPI_ISL_3185194, EPI_ISL_3185195, EPI_ISL_3185196, EPI_ISL_3185197, EPI_ISL_3185210, EPI_ISL_3185235, EPI_ISL_3185236, EPI_ISL_3185237, EPI_ISL_3185238, EPI_ISL_3185239, EPI_ISL_3185240, EPI_ISL_3185250, EPI_ISL_3185251, EPI_ISL_3186079, EPI_ISL_3186083, EPI_ISL_3186107, EPI_ISL_3186108, EPI_ISL_3186109, EPI_ISL_3186110, EPI_ISL_3186111, EPI_ISL_3186112, EPI_ISL_3186113, EPI_ISL_3186114, EPI_ISL_3186115, EPI_ISL_3186116, EPI_ISL_3186117, EPI_ISL_3186118, EPI_ISL_3186119, EPI_ISL_3186120, EPI_ISL_3186121, EPI_ISL_3186122, EPI_ISL_3186123, EPI_ISL_3186124, EPI_ISL_3186125, EPI_ISL_3186126, EPI_ISL_3186127, EPI_ISL_3186128, EPI_ISL_3186129, EPI_ISL_3186130, EPI_ISL_3186131, EPI_ISL_3186132, EPI_ISL_3186133, EPI_ISL_3186134, EPI_ISL_3186135, EPI_ISL_3186136, EPI_ISL_3186137, EPI_ISL_3186138, EPI_ISL_3186139, EPI_ISL_3186140, EPI_ISL_3186141, EPI_ISL_3186142, EPI_ISL_3186143, EPI_ISL_3186144, EPI_ISL_3186145, EPI_ISL_3186146, EPI_ISL_3186147, EPI_ISL_3186148, EPI_ISL_3186149, EPI_ISL_3186150, EPI_ISL_3186151, EPI_ISL_3186152, EPI_ISL_3186153, EPI_ISL_3186154, EPI_ISL_3186155, EPI_ISL_3186156, EPI_ISL_3186157, EPI_ISL_3186158, EPI_ISL_3186159, EPI_ISL_3186160, EPI_ISL_3186161, EPI_ISL_3186162, EPI_ISL_3186163, EPI_ISL_3186164, EPI_ISL_3186165, EPI_ISL_3186166, EPI_ISL_3186167, EPI_ISL_3186168, EPI_ISL_3186169, EPI_ISL_3186170, EPI_ISL_3186171, EPI_ISL_3186172, EPI_ISL_3186173, EPI_ISL_3186174, EPI_ISL_3186175, EPI_ISL_3186189, EPI_ISL_3186190, EPI_ISL_3186191, EPI_ISL_3186192, EPI_ISL_3186193, EPI_ISL_3186194, EPI_ISL_3186195, EPI_ISL_3186196, EPI_ISL_3186197, EPI_ISL_3186198, EPI_ISL_3186199, EPI_ISL_3186200, EPI_ISL_3186201, EPI_ISL_3186202, EPI_ISL_3186227, EPI_ISL_3186275, EPI_ISL_3186580, EPI_ISL_3186581, EPI_ISL_3186582, EPI_ISL_3186583, EPI_ISL_3186584, EPI_ISL_3186614, EPI_ISL_3186615, EPI_ISL_3186616, EPI_ISL_3186617, EPI_ISL_3186618, EPI_ISL_3186619, EPI_ISL_3186620, EPI_ISL_3186621, EPI_ISL_3186622, EPI_ISL_3186623, EPI_ISL_3186680, EPI_ISL_3186681, EPI_ISL_3186682, EPI_ISL_3186683, EPI_ISL_3186684, EPI_ISL_3186685, EPI_ISL_3186686, EPI_ISL_3186715, EPI_ISL_3186716, EPI_ISL_3186717, EPI_ISL_3186718, EPI_ISL_3186751, EPI_ISL_3186914, EPI_ISL_3186915, EPI_ISL_3186916, EPI_ISL_3186917, EPI_ISL_3186918, EPI_ISL_3186919, EPI_ISL_3186920, EPI_ISL_3186921, EPI_ISL_3186922, EPI_ISL_3186923, EPI_ISL_3186924, EPI_ISL_3186925, EPI_ISL_3186926, EPI_ISL_3186927, EPI_ISL_3186928, EPI_ISL_3186929, EPI_ISL_3186930, EPI_ISL_3186931, EPI_ISL_3186932, EPI_ISL_3186999, EPI_ISL_3187000, EPI_ISL_3187001, EPI_ISL_3187002, EPI_ISL_3187003, EPI_ISL_3187004, EPI_ISL_3187005, EPI_ISL_3187006, EPI_ISL_3187007, EPI_ISL_3187008, EPI_ISL_3187009, EPI_ISL_3187010, EPI_ISL_3187011, EPI_ISL_3187012, EPI_ISL_3187013, EPI_ISL_3187014, EPI_ISL_3187015, EPI_ISL_3187016, EPI_ISL_3187104, EPI_ISL_3187105, EPI_ISL_3187106, EPI_ISL_3187107, EPI_ISL_3187108, EPI_ISL_3187198, EPI_ISL_3187199, EPI_ISL_3187201, EPI_ISL_3187202, EPI_ISL_3187221, EPI_ISL_3187223, EPI_ISL_3187224, EPI_ISL_3187228, EPI_ISL_3281017, EPI_ISL_3281018, EPI_ISL_3281029, EPI_ISL_3281030, EPI_ISL_3281050, EPI_ISL_3305462, EPI_ISL_3305474, EPI_ISL_3305507, EPI_ISL_3305509, EPI_ISL_3305511, EPI_ISL_3305513, EPI_ISL_3398388, EPI_ISL_3398398, EPI_ISL_3398399, EPI_ISL_3398400, EPI_ISL_3398401, EPI_ISL_3398406, EPI_ISL_3398410, EPI_ISL_3398411, EPI_ISL_3398440, EPI_ISL_3398441, EPI_ISL_3398442, EPI_ISL_3398443, EPI_ISL_3398444, EPI_ISL_3398445, EPI_ISL_3398446, EPI_ISL_3398447, EPI_ISL_3398448, EPI_ISL_3398449, EPI_ISL_3398450, EPI_ISL_3398462, EPI_ISL_3398463, EPI_ISL_3398464, EPI_ISL_3398465, EPI_ISL_3398466, EPI_ISL_3398467, EPI_ISL_3398468, EPI_ISL_3398469, EPI_ISL_3398470, EPI_ISL_3398471, EPI_ISL_3398472, EPI_ISL_3398486, EPI_ISL_3398487, EPI_ISL_3398488, EPI_ISL_3398501, EPI_ISL_3398542, EPI_ISL_3398548, EPI_ISL_3398556, EPI_ISL_3398565, EPI_ISL_3398599, EPI_ISL_3398600, EPI_ISL_3398601, EPI_ISL_3398618, EPI_ISL_3398619, EPI_ISL_3398620, EPI_ISL_3398621, EPI_ISL_3398717, EPI_ISL_3398718, EPI_ISL_3398719, EPI_ISL_3398720, EPI_ISL_3398721, EPI_ISL_3398722, EPI_ISL_3398723, EPI_ISL_3398724, EPI_ISL_3398725, EPI_ISL_3398726, EPI_ISL_3401753, EPI_ISL_3401754, EPI_ISL_3401755, EPI_ISL_3401756, EPI_ISL_3401757, EPI_ISL_3401758, EPI_ISL_3401759, EPI_ISL_3401760, EPI_ISL_3401761, EPI_ISL_3401762, EPI_ISL_3401763, EPI_ISL_3401764, EPI_ISL_3401765, EPI_ISL_3401766, EPI_ISL_3401767 | | | |
| see above | St Vincent's Pathology (SydPath) | NSW Health Pathology - Institute of Clinical Pathology and Medical Research; Westmead Hospital; University of Sydney | Arnott A.; CIDM-PH et al.; Draper J.; Gall M.; Martinez E.; Rockett R.; Sintchenko V.; on behalf of ICPMR |
| EPI_ISL_912186, EPI_ISL_3185139, EPI_ISL_3185199, EPI_ISL_3185200, EPI_ISL_3186671, EPI_ISL_3186989, EPI_ISL_3281037, EPI_ISL_3281038, EPI_ISL_3281137, EPI_ISL_3281138, EPI_ISL_3281233, EPI_ISL_3281234, EPI_ISL_3281235, EPI_ISL_3281338, EPI_ISL_3281339, EPI_ISL_3281340, EPI_ISL_3398405 | | | |
| see above | Sydney South West Pathology Service (SSWPS) - Concord Repatriation General Hospital - NSW Health Pathology | NSW Health Pathology - Institute of Clinical Pathology and Medical Research; Westmead Hospital; University of Sydney | Arnott A.; CIDM-PH et al.; Draper J.; Gall M.; Martinez E.; Rockett R.; Sintchenko V.; on behalf of ICPMR |
| EPI_ISL_767939, EPI_ISL_767940, EPI_ISL_806728, EPI_ISL_872578, EPI_ISL_2811986, EPI_ISL_2811987, EPI_ISL_2811988, EPI_ISL_2812821, EPI_ISL_2812822, EPI_ISL_2812827, EPI_ISL_2812831, EPI_ISL_2812832, EPI_ISL_2812833, EPI_ISL_2812834, EPI_ISL_2812835, EPI_ISL_2812836, EPI_ISL_2812837, EPI_ISL_2812838, EPI_ISL_2812839, EPI_ISL_2812840, EPI_ISL_2812841, EPI_ISL_2812855, EPI_ISL_2812857, EPI_ISL_2812872, EPI_ISL_2812877, EPI_ISL_2812896, EPI_ISL_2812897, EPI_ISL_2828079, EPI_ISL_2828085, EPI_ISL_2828087, EPI_ISL_2828088, EPI_ISL_2828089, EPI_ISL_2861256, EPI_ISL_2876043, EPI_ISL_2876044, EPI_ISL_2876045, EPI_ISL_2894141, EPI_ISL_2894142, EPI_ISL_3007281, EPI_ISL_3007282, EPI_ISL_3007283, EPI_ISL_3007284, EPI_ISL_3007285, EPI_ISL_3007286, EPI_ISL_3007287, EPI_ISL_3007288, EPI_ISL_3007294, EPI_ISL_3007301, EPI_ISL_3015158, EPI_ISL_3015159, EPI_ISL_3015160, EPI_ISL_3015161, EPI_ISL_3015162, EPI_ISL_3119464, EPI_ISL_3184882, EPI_ISL_3184887, EPI_ISL_3184895, EPI_ISL_3184904, EPI_ISL_3184911, EPI_ISL_3184917, EPI_ISL_3184922, EPI_ISL_3184925, EPI_ISL_3184929, EPI_ISL_3184933, EPI_ISL_3185056, EPI_ISL_3185057, EPI_ISL_3185064, EPI_ISL_3185065, EPI_ISL_3185066, EPI_ISL_3185067, EPI_ISL_3185068, EPI_ISL_3185069, EPI_ISL_3185070, EPI_ISL_3185071, EPI_ISL_3185072, EPI_ISL_3185073, EPI_ISL_3185136, EPI_ISL_3185137, EPI_ISL_3185138, EPI_ISL_3185201, EPI_ISL_3185202, EPI_ISL_3185203, EPI_ISL_3185234, EPI_ISL_3185241, EPI_ISL_3185242, EPI_ISL_3185253, EPI_ISL_3185254, EPI_ISL_3185255, EPI_ISL_3185256, EPI_ISL_3186078, EPI_ISL_3186081, EPI_ISL_3186094, EPI_ISL_3186095, EPI_ISL_3186096, EPI_ISL_3186097, EPI_ISL_3186098, EPI_ISL_3186099, EPI_ISL_3186100, EPI_ISL_3186101, EPI_ISL_3186102, EPI_ISL_3186103, EPI_ISL_3186104, EPI_ISL_3186105, EPI_ISL_3186106, EPI_ISL_3186185, EPI_ISL_3186186, EPI_ISL_3186187, EPI_ISL_3186188, EPI_ISL_3186272, EPI_ISL_3186274, EPI_ISL_3186472, EPI_ISL_3186473, EPI_ISL_3186474, EPI_ISL_3186475, EPI_ISL_3186476, EPI_ISL_3186477, EPI_ISL_3186478, EPI_ISL_3186479, EPI_ISL_3186480, EPI_ISL_3186481, EPI_ISL_3186482, EPI_ISL_3186483, EPI_ISL_3186488, EPI_ISL_3186489, EPI_ISL_3186579, EPI_ISL_3186606, EPI_ISL_3186607, EPI_ISL_3186608, EPI_ISL_3186609, EPI_ISL_3186610, EPI_ISL_3186611, EPI_ISL_3186612, EPI_ISL_3186676, EPI_ISL_3186677, EPI_ISL_3186698, EPI_ISL_3186699, EPI_ISL_3186700, EPI_ISL_3186701, EPI_ISL_3186702, EPI_ISL_3186703, EPI_ISL_3186704, EPI_ISL_3186743, EPI_ISL_3186744, EPI_ISL_3186802, EPI_ISL_3186898, EPI_ISL_3186899, EPI_ISL_3186900, EPI_ISL_3186901, EPI_ISL_3186902, EPI_ISL_3186903, EPI_ISL_3186973, EPI_ISL_3186974, EPI_ISL_3186975, EPI_ISL_3186976, EPI_ISL_3186977, EPI_ISL_3186978, EPI_ISL_3186979, EPI_ISL_3186980, EPI_ISL_3186981, EPI_ISL_3186982, EPI_ISL_3186983, EPI_ISL_3186984, EPI_ISL_3186985, EPI_ISL_3186986, EPI_ISL_3186987, EPI_ISL_3186988, EPI_ISL_3186990, EPI_ISL_3186991, EPI_ISL_3186992, EPI_ISL_3186993, EPI_ISL_3186994, EPI_ISL_3186995, EPI_ISL_3187090, EPI_ISL_3187091, EPI_ISL_3187092, EPI_ISL_3187093, EPI_ISL_3187152, EPI_ISL_3187153, EPI_ISL_3187154, EPI_ISL_3187155, EPI_ISL_3187156, EPI_ISL_3187157, EPI_ISL_3187158, EPI_ISL_3187160, EPI_ISL_3187161, EPI_ISL_3187162, EPI_ISL_3187163, EPI_ISL_3187164, EPI_ISL_3187165, EPI_ISL_3281027, EPI_ISL_3281032, EPI_ISL_3281047, EPI_ISL_3281048, EPI_ISL_3281049, EPI_ISL_3281056, EPI_ISL_3305413, EPI_ISL_3305416, EPI_ISL_3305421, EPI_ISL_3305428, EPI_ISL_3305429, EPI_ISL_3305431, EPI_ISL_3305437, EPI_ISL_3305439, EPI_ISL_3305443, EPI_ISL_3305457, EPI_ISL_3305459, EPI_ISL_3305460, EPI_ISL_3305467, EPI_ISL_3305471, EPI_ISL_3305485, EPI_ISL_3305487, EPI_ISL_3305489, EPI_ISL_3398394, EPI_ISL_3398395, EPI_ISL_3398396, EPI_ISL_3398397, EPI_ISL_3398407, EPI_ISL_3398427, EPI_ISL_3398428, EPI_ISL_3398429, EPI_ISL_3398459, EPI_ISL_3398460, EPI_ISL_3398461, EPI_ISL_3398496, EPI_ISL_3398497, EPI_ISL_3398498, EPI_ISL_3398499, EPI_ISL_3398500, EPI_ISL_3398538, EPI_ISL_3398539, EPI_ISL_3398540, EPI_ISL_3398541, EPI_ISL_3398558, EPI_ISL_3398563, EPI_ISL_3398564, EPI_ISL_3398597, EPI_ISL_3398598, EPI_ISL_3398649, EPI_ISL_3398650, EPI_ISL_3398651, EPI_ISL_3401717, EPI_ISL_3401718, | | | |

| | | | |
|---|---|---|---|
| EPI_ISL_3401719, EPI_ISL_3401720, EPI_ISL_3401721, EPI_ISL_3401722, EPI_ISL_3401723, EPI_ISL_3401724, EPI_ISL_3401725, EPI_ISL_3401726, EPI_ISL_3401727, EPI_ISL_3401728, EPI_ISL_3401729, EPI_ISL_3401730, EPI_ISL_3401731, EPI_ISL_3401732, EPI_ISL_3401733, EPI_ISL_3401734, EPI_ISL_3401735, EPI_ISL_3401736, EPI_ISL_3401737, EPI_ISL_3401738, EPI_ISL_3401739, EPI_ISL_3401751, EPI_ISL_3401752, EPI_ISL_3401783, EPI_ISL_3401786, EPI_ISL_3401790, EPI_ISL_3401791, EPI_ISL_3401792 | | | |
| see above | Sydney South West Pathology Service (SSWPS) - Liverpool Hospital - NSW Health Pathology | NSW Health Pathology - Institute of Clinical Pathology and Medical Research; Westmead Hospital; University of Sydney | Arnott A.; CIDM-PH et al.; Draper J.; Gall M.; Martinez E.; Rockett R.; Sintchenko V.; on behalf of ICPMR |
| EPI_ISL_803110, EPI_ISL_803111, EPI_ISL_803112, EPI_ISL_803113, EPI_ISL_803114, EPI_ISL_806729, EPI_ISL_825016, EPI_ISL_845798, EPI_ISL_845799, EPI_ISL_845805, EPI_ISL_845806, EPI_ISL_872579, EPI_ISL_872586, EPI_ISL_872587, EPI_ISL_872588, EPI_ISL_914875, EPI_ISL_914876, EPI_ISL_914877, EPI_ISL_1383246, EPI_ISL_1904446, EPI_ISL_1904447, EPI_ISL_1904448, EPI_ISL_1904467, EPI_ISL_2405111, EPI_ISL_2405112, EPI_ISL_2462409, EPI_ISL_2462411, EPI_ISL_2462415, EPI_ISL_2462417, EPI_ISL_2462421, EPI_ISL_2462426, EPI_ISL_2462427, EPI_ISL_2462428, EPI_ISL_2462429, EPI_ISL_2650468, EPI_ISL_2650469, EPI_ISL_2650470, EPI_ISL_2828059, EPI_ISL_2828060, EPI_ISL_2828061, EPI_ISL_2828062, EPI_ISL_2828063, EPI_ISL_2828064, EPI_ISL_2828065, EPI_ISL_2828066, EPI_ISL_2828067, EPI_ISL_2828068, EPI_ISL_3184935, EPI_ISL_3184973, EPI_ISL_3184974, EPI_ISL_3184975, EPI_ISL_3184976, EPI_ISL_3184977, EPI_ISL_3184978, EPI_ISL_3184979, EPI_ISL_3184980, EPI_ISL_3184981, EPI_ISL_3184982, EPI_ISL_3184983, EPI_ISL_3184984, EPI_ISL_3185003, EPI_ISL_3185004, EPI_ISL_3185005, EPI_ISL_3185006, EPI_ISL_3185007, EPI_ISL_3185040, EPI_ISL_3185041, EPI_ISL_3185042, EPI_ISL_3185043, EPI_ISL_3185213, EPI_ISL_3185214, EPI_ISL_3185215, EPI_ISL_3185216, EPI_ISL_3185217, EPI_ISL_3185218, EPI_ISL_3185219, EPI_ISL_3185220, EPI_ISL_3185221, EPI_ISL_3185222, EPI_ISL_3185224, EPI_ISL_3185225, EPI_ISL_3185226, EPI_ISL_3185233, EPI_ISL_3185258, EPI_ISL_3305411, EPI_ISL_3398381, EPI_ISL_3398382, EPI_ISL_3398383 | | | |
| see above | Sydney South West Pathology Service (SSWPS) - Royal Prince Alfred Hospital - NSW Health Pathology | NSW Health Pathology - Institute of Clinical Pathology and Medical Research; Westmead Hospital; University of Sydney | Arnott A.; CIDM-PH et al.; Draper J.; Gall M.; Martinez E.; Rockett R.; Sintchenko V.; on behalf of ICPMR |
| EPI_ISL_3185198, EPI_ISL_3185252, EPI_ISL_3186466, EPI_ISL_3186467, EPI_ISL_3186591, EPI_ISL_3186678, EPI_ISL_3186679, EPI_ISL_3186748, EPI_ISL_3186752, EPI_ISL_3186756, EPI_ISL_3186904, EPI_ISL_3186905, EPI_ISL_3186906, EPI_ISL_3186907, EPI_ISL_3186908, EPI_ISL_3186909, EPI_ISL_3187175, EPI_ISL_3187211, EPI_ISL_3187212, EPI_ISL_3187225, EPI_ISL_3281306, EPI_ISL_3281307, EPI_ISL_3281308, EPI_ISL_3281309, EPI_ISL_3281310, EPI_ISL_3281311, EPI_ISL_3281312, EPI_ISL_3281313, EPI_ISL_3281314, EPI_ISL_3281315, EPI_ISL_3281316, EPI_ISL_3281317, EPI_ISL_3281318, EPI_ISL_3281319, EPI_ISL_3281320, EPI_ISL_3281321, EPI_ISL_3281322, EPI_ISL_3398534, EPI_ISL_3398535, EPI_ISL_3398557, EPI_ISL_3398659 | | | |
| see above | The Children's Hospital at Westmead | NSW Health Pathology - Institute of Clinical Pathology and Medical Research; Westmead Hospital; University of Sydney | Arnott A.; CIDM-PH et al.; Draper J.; Gall M.; Martinez E.; Rockett R.; Sintchenko V.; on behalf of ICPMR |
| EPI_ISL_779648, EPI_ISL_779649, EPI_ISL_812435, EPI_ISL_812436, EPI_ISL_812438, EPI_ISL_812439, EPI_ISL_812440, EPI_ISL_812441, EPI_ISL_812442, EPI_ISL_854751, EPI_ISL_854752, EPI_ISL_854753, EPI_ISL_854757, EPI_ISL_854759, EPI_ISL_854762, EPI_ISL_877564, EPI_ISL_877565, EPI_ISL_877566, EPI_ISL_877567, EPI_ISL_877568, EPI_ISL_877569, EPI_ISL_877576, EPI_ISL_877577, EPI_ISL_877578, EPI_ISL_877582, EPI_ISL_877583, EPI_ISL_877584, EPI_ISL_933777, EPI_ISL_933779, EPI_ISL_962823, EPI_ISL_962824, EPI_ISL_962825, EPI_ISL_962828, EPI_ISL_962829, EPI_ISL_962830, EPI_ISL_962831, EPI_ISL_962832, EPI_ISL_979356, EPI_ISL_979362, EPI_ISL_979363, EPI_ISL_979364, EPI_ISL_1033154, EPI_ISL_1033155, EPI_ISL_1249993, EPI_ISL_1249996, EPI_ISL_1913198, EPI_ISL_1913199, EPI_ISL_1913200, EPI_ISL_1913201, EPI_ISL_1913202, EPI_ISL_1913203, EPI_ISL_1913204, EPI_ISL_1913205, EPI_ISL_1913206, EPI_ISL_1913207, EPI_ISL_1913208, EPI_ISL_1913209, EPI_ISL_1913211, EPI_ISL_1913212, EPI_ISL_2920925, EPI_ISL_2920927, EPI_ISL_2920928, EPI_ISL_2920929, EPI_ISL_2920930, EPI_ISL_2920931, EPI_ISL_2920932, EPI_ISL_2920933, EPI_ISL_2920934, EPI_ISL_2920935, EPI_ISL_2920936, EPI_ISL_2920937, EPI_ISL_2920938, EPI_ISL_2920939, EPI_ISL_2920940, EPI_ISL_2920941, EPI_ISL_2920942, EPI_ISL_2920943, EPI_ISL_2920944, EPI_ISL_2920945, EPI_ISL_2920946, EPI_ISL_2920947, EPI_ISL_2920948, EPI_ISL_2920949, EPI_ISL_2920950, EPI_ISL_2920951, EPI_ISL_2920952, EPI_ISL_2920953, EPI_ISL_2920954, EPI_ISL_2920955, EPI_ISL_2920956, EPI_ISL_2920957, EPI_ISL_2920958, EPI_ISL_2920959, EPI_ISL_2920960, EPI_ISL_2920961, EPI_ISL_2920962, EPI_ISL_2920963, EPI_ISL_2920964, EPI_ISL_2920965, EPI_ISL_2920966, EPI_ISL_2920967, EPI_ISL_2920968, EPI_ISL_2920969, EPI_ISL_2920970, EPI_ISL_2920971, EPI_ISL_2920972, EPI_ISL_2920973, EPI_ISL_2920974, EPI_ISL_2920975, EPI_ISL_2920976, EPI_ISL_2920977, EPI_ISL_2920978, EPI_ISL_2920979, EPI_ISL_2920980, EPI_ISL_2920981, EPI_ISL_2920982, EPI_ISL_2920983, EPI_ISL_2920984, EPI_ISL_2920985, EPI_ISL_2920986, EPI_ISL_2920987, EPI_ISL_2920988, EPI_ISL_2920989, EPI_ISL_2920990, EPI_ISL_2920991, EPI_ISL_2920992, EPI_ISL_2920993, EPI_ISL_2920994, EPI_ISL_2920995, EPI_ISL_2920996, EPI_ISL_2920997, EPI_ISL_2920998, EPI_ISL_2920999, EPI_ISL_2921000, EPI_ISL_2978348, EPI_ISL_2978349, EPI_ISL_2978350, EPI_ISL_2978351, EPI_ISL_3030411, EPI_ISL_3030414, EPI_ISL_3030415, EPI_ISL_3030416, EPI_ISL_3050801, EPI_ISL_3134919, EPI_ISL_3134920 | | | |
| see above | Victorian Infectious Diseases Reference Laboratory (VIDRL) | VIDRL and MDU-PHL | Caly L.; Druce J.; M.L.; N.L.; Sait; Seemann T.; Sherry |
| EPI_ISL_1938308, EPI_ISL_2675251 | unknown | PHV-FSS | Son Nguyen |
